# A Human-in-the-Loop Large Language Model System Based on the Model Context Protocol for Differential Diagnosis from Electronic Medical Records and Literature

**DOI:** 10.64898/2026.08.18.26359085

**Authors:** Hyunseok Lim, Junyoung Yoon, Hahn Yi, Heeyeon Kwon, Dong-Wook Lee, Namkug Kim

## Abstract

Diagnostic errors, including misdiagnoses and delayed clinical diagnoses, could affect outcomes of a significant patient population, particularly individuals presenting with rare diseases or non-specific symptoms. From rule-based diagnostic decision supporting systems (DDSS) to large language model (LLM) based tools for clinical reasoning have been developed to address these limitations. However, existing DDSS are often proprietary and difficult to integrate, and recent LLM-based tools remain hindered by operational challenges such as cost, resources constraint, and privacy concerns. Moreover, existing systems interpret electronic medical records (EMR) and generate diagnoses separately, limiting continuous evidence-based analysis and imposing repeated clinician involvement. In this paper, we present DDx-Finder, an open-source framework that leverages Model Context Protocol (MCP) servers for direct EMR and literature access, enabling prompt-driven clinical state extraction and reliable case-report retrieval via generating searching query by LLM, while addressing limitations related to resource demands and privacy concerns. A clinical case study demonstrates the system’s feasibility and its potential to provide accessible, transparent, and systematic differential diagnostic support for complex cases. The implementation is available at https://github.com/loopback-kr/DDx-Finder.

## 1 Introduction

Diagnostic errors pose a significant challenge in healthcare, with a study estimating that 10–15% of clinical diagnoses may be incorrect or delayed, affecting millions of patients annually [62]. These errors are particularly prevalent in the initial diagnosis of rare diseases and in chronically ill patients presenting with non-specific symptoms, as both scenarios inherently complicate the generation of comprehensive differential diagnoses (DDx) [11, 26]. A critical contributing factor is the failure to systematically consider relevant DDx during clinical evaluation[29]. Given the pervasive nature and serious consequences of these errors, improving diagnostic processes—particularly supporting clinicians in differential diagnosis generation—has become a healthcare priority[9].

Clinicians face substantial practical barriers when attempting to systematically consider DDx in routine practice. Electronic medical records (EMRs), while essential, impose significant cognitive burdens on providers. The mental effort required to navigate fragmented patient data, identify relevant information, and maintain diagnostic reasoning increases cognitive workload substantially [54, 6]. Extended EMR interaction time is strongly associated with clinician burnout [45]. Beyond the challenges withing the EMR, retrieving relevant medical literature from databases and comprehending their clinical content requires specialized medical expertise and consumes considerable clinical time[21, 1, 66]. These compounded challenges often lead clinicians to rely on readily available information sources rather than comprehensive evidence searches to support DDx generation, particularly under time pressure[4].

To address diagnostic errors, clinical decision support systems (CDSS) have been widely deployed to improve care quality, primarily through real-time alerts, medication safety checks, and protocol enforcement[64]. While effective for routine clinical workflows, these systems have shown limited utility in supporting differential diagnosis generation for complex or ambiguous cases—particularly those involving rare diseases or atypical presentations of chronic conditions [29, 11, 26]. Such cases require iterative hypothesis generation and synthesis of diverse clinical evidence, including analysis of similar cases from medical literature [25, 18].

Various computational approaches have been developed over several decades to support differential diagnosis generation. Rule-based diagnostic support systems have been developed over several decades, encoding expert medical knowledge into computational frameworks[60, 48]. While more recent systems have improved accessibility through web-based interfaces, expanded knowledge bases, and incorporated machine learning approaches[30, 53], effective integration of structured expert knowledge with data-driven methods and maintenance of evolving medical knowledge remain challenges[13, 49].

Recent AI systems employing large language model (LLM) architectures have shown promising diagnostic capabilities across clinical reasoning tasks, with advances in both single-model approaches and multi-agent frameworks[27, 59, 27, 59, 14, 44, 56, 20]. However, these systems face deployment barriers including cost, privacy concerns, limited accessibility of proprietary APIs, and reliability issues such as hallucinations and semantic drift[58, 63, 15, 70]. Beyond these practical challenges, a fundamental architectural limitation is that most treat EMR data ingestion and diagnostic synthesis as separate stages, limiting iterative, evidence-grounded reasoning workflows essential for complex diagnostic cases. There is thus a need for an integrated, open-source framework that enables direct EMR access with human-in-the-loop DDx support to mitigate LLM hallucinations and semantic drift—limitations that currently necessitate human oversight in complex clinical contexts.

We present DDx-Finder, an open-source framework that addresses these limitations through two integrated capabilities: (1) automated extraction of patient clinical status and ruled-out diagnoses from EMR data with evidence linking, and (2) generation of targeted case report queries and synthesis of retrieved literature based on the extracted clinical context. By leveraging open-source LLM components, our modular architecture offers a reproducible, costeffective alternative to proprietary solutions. We review related approaches in Section 2, detail our system design in Section 3, validate feasibility through clinical case studies in Section 4, and discuss implications in Section 5.

**Contributions.** This work makes the following contributions:

- A practical modular architecture that integrates direct EMR access with LLM-based diagnostic support using the Model Context Protocol (MCP), an open standard for connecting LLMs to external tools
- An open-source implementation deployable in security-conscious healthcare institutions
- Structured methods for clinical state extraction and ruled-out diagnosis identification using simple user prompts without complex system prompts or complex Prompt Engineering techniques, enabling seamless integration into clinical workflows
- A proof-of-concept demonstration on a diagnostically complex clinical case

## 2 Related Work

Building on the diagnostic challenges outlined in Section 1, we review the evolution of AI-assisted diagnosis from early knowledge-based systems through modern LLM-based approaches to agentic, tool-mediated systems that interact directly with clinical data.

### 2.1 Diagnostic Decision Support: From Rule-Based to Data-Driven

Rule-based diagnostic support systems emerged in the 1970s-80s, encoding expert medical knowledge into computational frameworks[60]. Early systems such as INTERNIST-1[47] and its successor QMR (Quick Medical Reference)[46] represented disease-symptom relationships through extensive knowledge bases covering thousands of conditions. During this period, DXplain[10, 24] employed modified Bayesian reasoning across 2,200+ diseases, while ILIAD[69] employed Bayesian networks to model probabilistic relationships in internal medicine, serving as both a clinical decision support tool and an educational platform for teaching differential diagnosis to students.

Web-based systems in the 2000s improved accessibility while maintaining knowledge-based foundations. Isabel[30], motivated by a misdiagnosis of the founder’s daughter, achieved commercial success as a web-based diagnostic tool employing pattern recognition for differential diagnosis generation. The system operated through both standalone symptom entry and EMR integration with natural language processing[36], also incorporating machine learning to expand beyond rule-based approaches. VisualDx[53] specialized in image-based diagnosis for dermatology and other visual specialties, enabling clinicians to search by entering patient descriptors and lesion morphologies rather than disease names [65]. The platform maintained a comprehensive image database and later incorporated DermExpert, a supervised learning-based diagnostic system trained on over 76,000 dermatologist-labeled images[23]. Both systems, however, remain proprietary platforms with limited transparency regarding their algorithmic approaches and ongoing development, restricting independent validation and reproducibility.

### 2.2 Medical LLMs and Diagnostic Reasoning

Recent advances in medical LLMs have shown that instruction-tuned systems can encode substantial clinical knowledge and support DDx reasoning. Early work such as Med-PaLM demonstrated that aligning models with clinician-style reasoning improves performance on medical QA and structured evaluation benchmarks[61]. Complementary assessments using published case reports further examined how general-purpose LLMs handle complex presentations, showing that diagnostic suggestions can be clinically useful yet sensitive to information completeness and prompt formulation[32]. Additional studies using clinicopathological conferences (CPC) case material explored methods for improving hypothesis structure and rationale generation, underscoring the importance of clearer intermediate reasoning processes in LLM-driven DDx tasks[44]. Beyond DDx, evidence-based platforms such as OpenEvidence examined how LLM-supported retrieval from trusted literature sources can assist primary-care decision-making, generally reinforcing rather than altering physician plans in common chronic conditions[33]. However, direct comparisons with established platforms suggest these advances have not yet translated to clear clinical superiority; ChatGPT-4 achieved 82.1% diagnostic accuracy compared to Isabel Pro’s 87.1% in a comparative evaluation[15], indicating that LLMs remain competitive with but not demonstrably superior to existing commercial diagnostic systems.

### 2.3 Agentic Approaches to Differential Diagnosis

Agentic approaches extend these foundations by exploring how LLMs can structure diagnostic work through iterative planning and targeted information acquisition. Systems such as MEDDxAgent exemplify this trend by decomposing DDx generation into a sequence of clarification, retrieval, and refinement steps that adjust as new clinical details are surfaced[56]. Multi-agent designs follow similar principles, with frameworks like RareAgents simulating role-based interactions to represent multidisciplinary diagnostic deliberation, particularly in settings where uncertainty and incomplete information are common[20]. Across these lines of work, the unifying trajectory is a shift toward interactive and tool-mediated agents. Across these lines of work, the unifying trajectory is a shift toward interactive and tool-mediated agents. However, these agentic frameworks remain in early stages of development, with limited evaluation in real-world clinical settings, necessitating rigorous validation before clinical deployment.

### 2.4 Clinical Tool Use and EMR Access

The Model Context Protocol (MCP), introduced by Anthropic as an open standard for connecting AI systems to external data sources and tools[5], provides a standardized framework for LLM-mediated access to clinical systems. Tool-use frameworks have begun to investigate how such agentic processes operate when connected to real clinical systems. The EHR-MCP framework evaluated LLM-mediated access to EMR and showed that models can reliably retrieve structured information through standardized MCP interfaces while revealing practical challenges in temporal queries, long-record handling, and tool-argument formulation[43]. Broader agentic clinical dicision-making systems similarly emphasize the need for controlled access to external resources to reduce hallucinations and maintain evidence grounding[2].

### 2.5 Literature Search

Point-of-care clinical information systems such as UpToDate and ClinicalKey have become widely adopted tools for evidence-based decision-making in practice settings. UpToDate, in particular, has demonstrated strong clinical preference due to its synthesis of recent literature into concise, actionable recommendations[38], with studies showing measurable impacts on clinical outcomes and bedside decision-making[34]. In contrast, ClinicalKey’s strength lies in comprehensive access to medical textbooks and journal databases, making it more suitable for didactic learning than rapid point-of-care queries[38]. More recently, AI-powered platforms such as OpenEvidence have emerged to provide rapid, evidence-based answers with transparent source citations, aiming to enhance usability for primarycare decision-making[33]. While these platforms provide curated evidence synthesis, they operate independently of patient-specific EMR data and require manual integration into the diagnostic workflow.

Despite this progress, many prior systems treat EMR ingestion and diagnostic synthesis as separate stages, relying either on pre-curated inputs or limited retrieval mechanisms. This leaves open questions about how diagnostic reasoning should operate when EMR access, query refinement, and literature search occur within a continuous workflow. Motivated by these gaps, the present work explores an approach that combines direct EMR access with agentic, stepwise diagnostic reasoning, aiming to support more coherent and clinically aligned DDx processes.

## 3 Methods and System Architecture

### 3.1 Overview

DDx-Finder consists of three main modules organized in a sequential pipeline (Figure 1):

1. **EMR Data Acquisition and Clinical State Extraction**: In a single user prompt, the LLM invokes the Filesystem MCP server’s summarize_medical_records tool to extract relevant EMR content in a token-efficient manner, then generates a clinical summary tailored to the user’s search objectives, typically including key symptoms, ruled-out diagnoses, and search-compatible terms. This initial output is reviewed by the user and can be iteratively refined through natural language prompts to adjust format, focus, or search strategy. Additional tools (get_full_lab_data, get_document_content) enable on-demand retrieval of complete records when needed.
2. **Query Generation and Human Refinement**: Based on the validated clinical summary, the LLM generates initial search queries using its pre-trained medical knowledge. The clinician must review and refine these queries through natural language instructions to ensure appropriate search strategy. Users can request query construction guidance through get_query_examples or get_pubmed_query_guide tools for useful examples and Boolean operators used to logically combine or exclude search keywords.
3. **Literature Retrieval**: The Medical Literature Search MCP server (search_literature tool) executes queries both PubMed and PMC databases, retrieving relevant case reports with title, abstract summaries, and direct links. When initial results are insufficient, the human reviewer evaluates the output, refines the query, and resubmits it, forming an iterative improvement loop.

**Figure 1:**
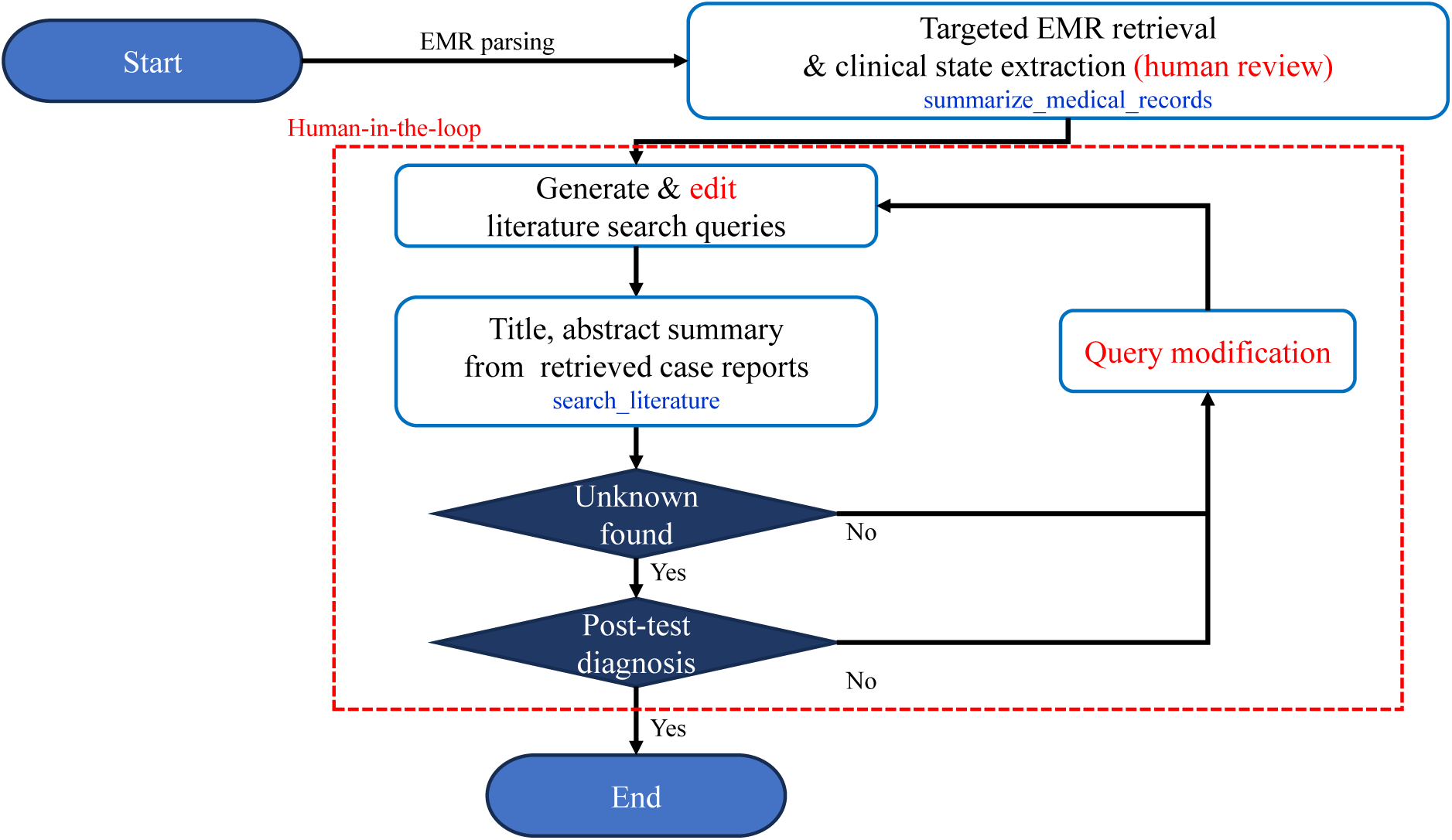
DDx-Finder workflow. DDx-Finder first parses EMR data and extracts clinical information using the summarize_medical_records tool, then generates literature-search queries. Clinicians review the extracted summary and refine the queries to improve retrieval. The search_literature tool returns titles, abstract summaries, and direct links to the top-k case reports. If no plausible unrecognized diagnosis is found, or if post-test evaluation does not support a candidate, clinicians modify the query and repeat the search until a clinically useful diagnosis is identified.

After retrieval, the clinician reviews the case reports to identify diagnostic candidates absent from the current differential diagnosis list. If no plausible candidate is found, the query is modified and the search is repeated. If a candidate is found but subsequent testing does not support the post-test diagnosis, the query is revised and another retrieval cycle is performed. The workflow emphasizes human oversight at critical junctions: after clinical state extraction and during query refinement, ensuring that automated suggestions are medically sound and search strategies are appropriately targeted. Additional implementation details—including model architectures, and server configurations—are provided in the Implementation Details.

### 3.2 EMR Acquisition and Clinical State Extraction

DDx-Finder performs EMR data retrieval and clinical summarization within a single user prompt through MCP tool-calling mechanism. When the practitioner issues a natural language instruction (e.g., “Review EMR data and summarize the patient’s current condition and exclusionary diagnoses based on imaging and lab test results, focusing on the reason for hospitalization rather than chronic background illness”), the following occurs in one interaction.

#### 3.2.1 Integrated Workflow

1. **Tool invocation**: The LLM recognizes the need for EMR access and automatically calls the summarize_ medical_records(start_date,end_date,top_n) tool via MCP
2. **Token-efficient data extraction**: The Filesystem MCP server applies rule-based filtering:
  - **Laboratory data**: Only abnormal findings are extracted based on visual markers—♦ (qualitative abnormal, score 100), ▲ (increased, score 10), and ▼ (decreased, score 10). By default, the top 15 most abnormal tests per date are returned (adjustable via top_n), reducing token consumption by about 20% while preserving clinically significant findings.
  - **Clinical notes**: Structural extraction targets diagnostically relevant sections—suspect diagnoses (pattern matching for “R/O”, “rule out”, max 10 items), chief complaint (200 characters), assessment (400 characters), and plan (300 characters).
3. **In-context analysis**: The extracted EMR data is inserted into the LLM’s context, which then generates a clinical summary without requiring separate prompting. The output typically includes:
  - Key symptoms and clinical findings with supporting evidence
  - Ruled-out diagnoses based on negative laboratory/imaging results
  - MeSH-compatible terminology for literature search

Critically, steps 1-3 occur within a single conversational turn. The user does not manually copy EMR data or issue separate analysis commands—the LLM autonomously orchestrates tool use and interpretation in response to the initial instruction. The detailed implementation algorithms are provided in Appendix A.

#### 3.2.2 Flexible Output Without Rigid Schema

No predefined JSON schema or output template is enforced. The LLM structures the summary based on its pretrained medical knowledge, with format and granularity varying by:

- **Model capability**: Summary structure and detail level vary with model size, with larger models providing more comprehensive organization
- **User instruction**: Natural language guidance (e.g., “focus on acute presentation”, “distinguish from chronic disease symptoms”) shapes the emphasis without system-level templates
- **Case complexity**: Multi-system presentations naturally yield more detailed categorization

This minimalist approach—relying solely on user prompts without system prompts or fine-tuning—ensures generalizability across diverse EMR formats, clinical scenarios, and institutional practices.

#### 3.2.3 Human-in-the-Loop Validation

The LLM-generated summary undergoes mandatory review before literature search:

1. **Medical accuracy**: Verifying correct interpretation of laboratory values, imaging findings, and temporal relationships
2. **Clinical focus refinement**: Adjusting emphasis via iterative prompts (e.g., reprioritizing findings, requesting reorganization by organ system)

Supplementary tools (get_full_lab_data(date), get_document_content(date,filename)) enable on-demand retrieval of complete records when the initial summary lacks specificity or when clinicians need to verify extracted data against source documents.

### 3.3 Query Generation and Human Refinement

Following validation of the clinical summary, the LLM generates initial literature search queries using its pre-trained medical knowledge to translate clinical presentations into database-appropriate search terms. These initial queries serve as starting points that clinicians iteratively refine through conversational instructions, adjusting the search strategy to broaden, narrow, or restructure queries based on their diagnostic reasoning.

When advanced query syntax is needed, two MCP tools provide guidance on request:

- get_query_examples(): Provides ready-to-use query templates with practical examples for databases (PubMed and PMC). Includes patterns for basic case report searches, exclusion criteria, and cross-database compatible syntax. Serves as a quick reference for common search scenarios.
- get_pubmed_query_guide(): Returns comprehensive PubMed-specific documentation for advanced users, covering field tag syntax ([Title/Abstract], [MeSH Terms]), Boolean operator precedence, MeSH term combinations, and advanced filters (date ranges, publication types). Designed for users requiring precise control over PubMed’s complex query features.

The LLM automatically invokes these tools when users request syntax help through natural language instructions, integrating the guidance into query formulation.

### 3.4 Literature Retrieval

The Medical Literature Search MCP server executes refined queries across two databases—PubMed (abstracts) and PMC (full-text articles)—through the search_literature tool. The system prioritizes query transparency and token efficiency while maintaining clinical utility.

#### 3.4.1 Multi-Database Search Execution

For each database, the tool:

1. Executes the user-specified query without modification or simplification
2. Retrieves up to a specified number of results per database (default: 20, configurable via max_results_per_db for uniform limits or max_results_by_db for database-specific limits)
3. Extracts query translation from each search engine showing how the query was interpreted
4. Parses XML responses to extract article metadata (title, authors, journal, year, abstract)

PubMed and PMC use NCBI’s E-utilities API with relevance-based sorting. Result ordering from each database is preserved to maintain search engine rankings.

#### 3.4.2 Token-Efficient Result Presentation

Retrieved articles are presented in compact tabular format with three columns:

- **Title**: Full article title with special character escaping for markdown compatibility
- **Summary**: First 70 words of the abstract (maximum 3 sentences), mechanically extracted. This word limit is configurable through the tool’s max_words parameter. Next, the LLM summarizes this extracted text according to the user’s instruction
- **Link**: Clickable identifier (PMID and PMC ID) linking to the full article

This format reduces token consumption by 60-80% compared to displaying complete abstracts while preserving sufficient information for relevance assessment. Duplicate articles (identified by title matching) are automatically removed.

#### 3.4.3 Query Transparency Features

The search_literature tool automatically extracts comprehensive search execution metadata and embeds it in the response structure. The tool returns results in one of three formats (controlled by return_format parameter):

- **Compact format** (default): Combines execution metadata and article table in a single content string
- **Detailed format**: Includes full abstracts with execution metadata
- **JSON format**: Separates execution metadata into a dedicated execution_summary field The execution metadata includes:
- **Executed query**: The exact query string sent to each database
- **Query translation**: How each search engine interpreted the query (e.g., automatic term mapping, field tag resolution), extracted from database API responses
- **Result count**: Number of articles retrieved per database
- **External search links**: Direct URLs to database search pages for exploring results beyond token limits

While always captured by the tool, metadata presentation is determined by the LLM based on user intent. Explicitly requesting execution details ensures their display. The complete literature search implementation is provided in Appendix B.

#### 3.4.4 Iterative Search Refinement

When initial results are insufficient or off-target, clinicians refine queries through direct manual editing or natural language instructions to the LLM. Manual refinement allows precise control—adding or removing terms, adjusting Boolean operators, or modifying field tags based on query transparency metadata from previous attempts. For instance, unexpected term mapping revealed in query translation can prompt explicit field tag usage. Conversational refinement through LLM instructions (e.g., “broaden the query by removing fever”) offers an alternative for users less familiar with database syntax. This iterative process continues until satisfactory case reports are identified.

### 3.5 Implementation Details

DDx-Finder is deployed on containerized software stacks hosted on a server equipped with GPUs, consisting of three primary containers that communicate through HTTP interfaces (Figure 2). Because the system processes sensitive EMR data that must not leave the hospital environment, all components handling patient information are deployed within an in-hospital on-premises system; only PubMed and PMC are accessed externally through the WAN for literature retrieval. We use gpt-oss as the core model — an open-source LLM that enables flexible customization and local deployment, in contrast to closed commercial LLMs. While several open-source LLMs such as Qwen, LLaMA, Mistral, and Falcon provide strong general-purpose capabilities, gpt-oss offers a more modular architecture[39], and strong compatibility with MCP-based tool systems[68], making it better suited for our application context. For model inference, we adopt vLLM[40], which provides high-throughput and memory-efficient serving compared to conventional Hugging Face Transformers or DeepSpeed inference setups. The user interface is built with Open WebUI[7], an extensible web-based framework that simplifies tool integration and multi-user management. To enable seamless integration between the model backend and tool execution layer, we employ MCPO (MCP-to-OpenAPI Proxy)[8], which serves as a lightweight connector bridging the gap between Open WebUI and the MCP Server. While the original MCP protocol relies on stdio communication — which lacks authentication, documentation, and standardized error handling — MCPO resolves these limitations by converting non-standard and insecure stdio streams into universally compatible HTTP/OpenAPI endpoints. From the clinician’s perspective, this integrated system enables interaction with LLMs and MCP servers through natural language alone. Detailed versions of each component are listed below:

- **Hardware**: Dual NVIDIA A100 GPUs (80GB VRAM each), Intel Xeon Gold 6226R CPU @ 2.90GHz (64 logical processors), 1TB RAM, Ubuntu 22.04 LTS
- **Serving LLM**: vLLM v0.10.2-x86_64 for gpt-oss-20b/120b serving
- **Model Configuration**: gpt-oss-120b with 32,768-token context window, representing the maximum window size supported by vLLM under the available GPU memory constraints
- **Web Interface**: Open WebUI v0.6.34 based on Node.js v22.21.1[50] for clinical interaction
- **MCP Integration**: FastMCP v2.12.4[41], MCPO v0.0.19 for EMR parsing and literature search MCP servers

**Figure 2:**
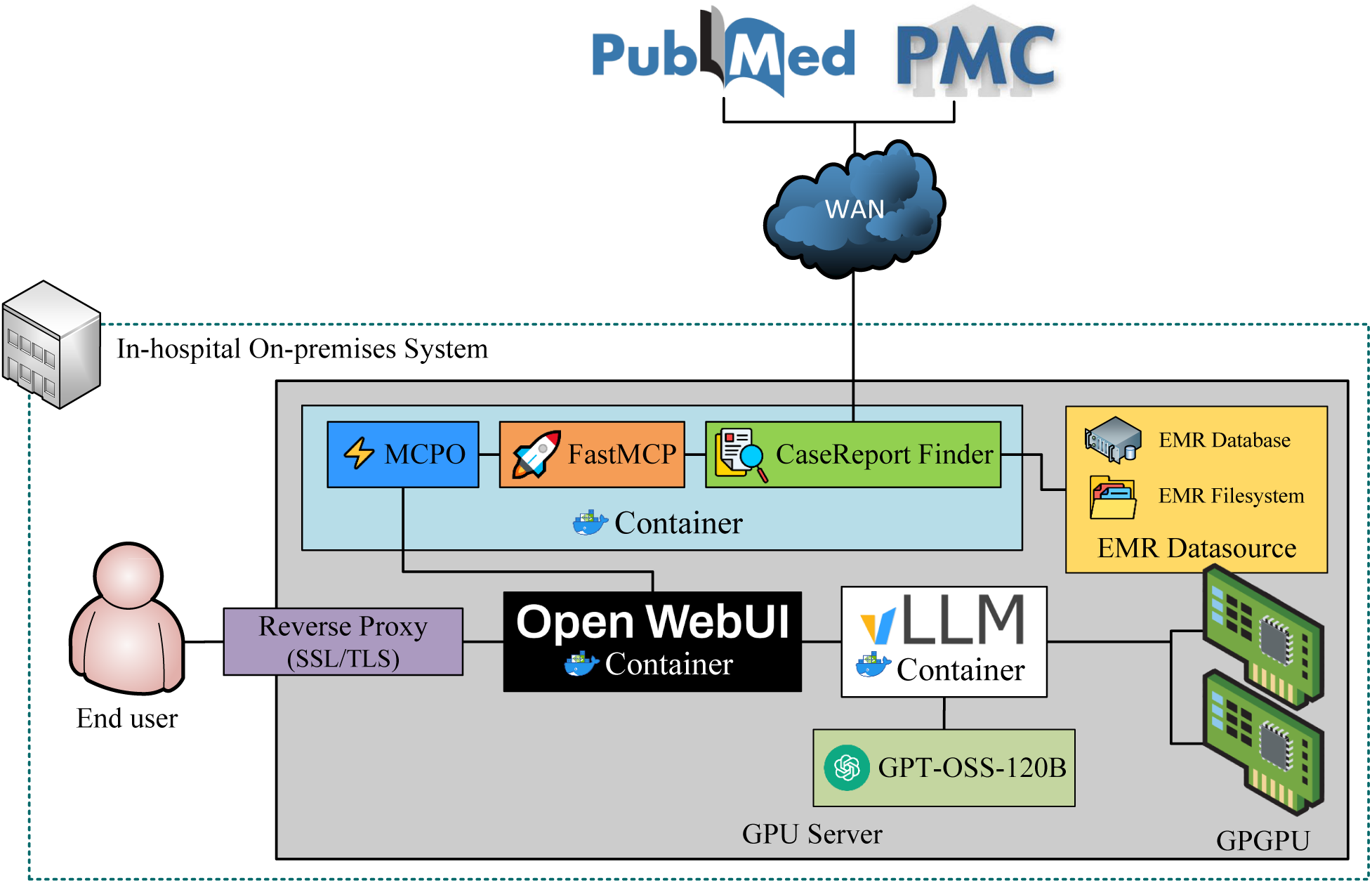
On-Premises DDx-Finder System Architecture for Secure In-Hospital EMR Processing

### 3.6 Ethical Considerations

This study evaluated a technological framework using a fully de-identified single clinical case for technical validation. As only anonymized retrospective data were used without patient risk, this study was determined to be exempt from formal review by the Institutional Review Board (IRB) of Asan Medical Center, Seoul, Korea (IRB Exemption No. 2026-1156) and was conducted in accordance with the ethical principles of the Declaration of Helsinki. The requirement for informed consent was waived by the IRB due to the retrospective nature of the study and the complete de-identification of the clinical data.

## 4 Case Study

### 4.1 Patient Presentation

We demonstrate DDx-Finder on an anonymized patient in their early 40s with end-stage renal disease on peritoneal dialysis presenting with:

- Bilateral thigh pain and elevated creatine kinase (5343 IU/L)
- Fatigue and cognitive slowing for the past several months
- Fragmented sleep due to ongoing diarrhea and bowel wall edema
- Elevated liver enzymes (AST 133, ALT 90 IU/L)
- Multiple electrolyte abnormalities with relatively prominent hyponatremia

### 4.2 Pipeline Execution

We apply DDx-Finder’s three-stage pipeline to this patient case using both gpt-oss-20b and gpt-oss-120b models to compare their clinical reasoning capabilities and proficiency in MCP tool usage.

#### Step 1: EMR Acquisition and Clinical State Extraction

The system invokes the Filesystem MCP server’s summarize_medical_records tool to retrieve summaries of 8 days of EMR data, including admission notes, progress notes, laboratory results, and imaging/procedural reports (chest X-ray, CT scan, echocardiogram, colonoscopy, esophagogastroduodenoscopy, and whole-body bone scan).

Both models generate clinical summaries identifying rhabdomyolysis, diarrhea, weakness, thigh pain, and worsening liver function. As shown in Table C.1, gpt-oss-120b produces more structured output with explicit MeSH-compatible terminology that accurately reflected context-specific symptoms. It adhered closely to user instructions emphasizing acute presentation over chronic ESRD symptoms, and identified a broader range of precise MeSH terms aligned with the patient’s hospitalization reason. In contrast, gpt-oss-20b (see Table C.3) occasionally generates contextually inappropriate MeSH mappings (e.g., “Femoral Pain” for thigh pain, “Insomnia” for sleep disturbance) and includes typical ESRD-related symptoms despite explicit instructions to focus on acute findings, requiring additional human refinement to achieve clinical accuracy.

#### Step 2: Ruled-Out Diagnoses Identification

Comparison of exclusion rationale between gpt-oss-20b and gpt-oss-120b (Tables C.4 and C.2)shows the following ruled-out differential diagnoses:

- Acute abdominal emergencies (appendicitis, mesenteric ischemia, bowel obstruction)
- Infectious etiologies (pyelonephritis/UTI, sepsis, acute viral hepatitis)
- Cardiovascular/vascular causes (AMI/CAD, pulmonary embolism, cardiac tamponade, vascular aneurysm)
- Hepatic and metabolic crises (severe hepatic failure, severe electrolyte derangement)
- Musculoskeletal causes (skeletal fractures)

In particular, although both models identify overlapping categories of excluded diagnoses across abdominal emergencies, infections, hepatic conditions, vascular disorders, and metabolic crises, their reasoning styles diverge distinctly: gpt-oss-20b emphasize abdominal and structural etiologies through predominantly qualitative, imaging-centered arguments, whereas gpt-oss-120b broaden the differential into cardiopulmonary and systemic internal medicine domains and support each exclusion with precise laboratory metrics, imaging interpretations, and pathophysiologic expectations. In short, similar to the clinical state extraction behavior, gpt-oss-120b appears to follow the instruction to focus on the patient’s acute presentation.

#### Step 3: Query Generation and Literature Retrieval

Based on validated clinical summaries, both models generate initial PubMed and PMC search queries with markedly different clinical reasoning approaches. As shown in Appendix C.2.2, gpt-oss-120b constructs a comprehensive query—(“Rhabdomyolysis”[Mesh] AND “Diarrhea”[Mesh]) AND (“Hyponatremia”[Mesh])—that prioritize frequently documented symptoms, clinically severe findings, and manifestations directly relevant to acute hospitalization rather than chronic ESRD sequelae. This query effectively captures the patient’s presenting constellation of muscle breakdown, gastrointestinal disturbance, and the most prominent electrolyte abnormality documented in laboratory results. gpt-oss-120b also proactively provides supplementary search guidance and query refinement strategies to enhance literature retrieval effectiveness. In contrast, gpt-oss-20b generates “Rhabdomyolysis”[Mesh] AND “Hyperkalemia”[Mesh], focusing on hyperkalemia—a life-threatening complication commonly associated with ESRD—while omitting hyponatremia despite its clinical prominence (see Appendix C.2.3). This reflects a tendency to emphasize chronic disease complications over acute presenting features, requiring iterative human refinement through natural language instructions to achieve clinically relevant queries.

To systematically compare model performance beyond query formulation, we execute identical search queries using both models and evaluated three key dimensions: (1) instruction adherence and accuracy in abstract summarization, (2) precision in matching retrieved case reports with direct access links, and (3) proficiency in MCP tool selection (see Appendix C.3). For abstract summarization, when both models generate 15-word summaries of a rhabdomyolysis case report, gpt-oss-120b accurately captures the core clinical message that AST/ALT elevation reflects muscle-derived enzymes rather than liver injury, aligning with the abstract’s intent to prevent diagnostic misinterpretation. gpt-oss20b, however, introduces a factual error by describing “suspected liver injury requiring hospitalization”—contradicting the original abstract’s emphasis that liver damage is *not* present—and added content absent from the source material. Regarding link accuracy, gpt-oss-120b generates correct direct links for all 20 case reports (10 PubMed, 10 PMC), where gpt-oss-20b produces one erroneous PubMed link by incorrectly listing a PMC-sourced case report in the PubMed table with a malformed URL, while PMC links remain accurate. For MCP tool selection, when prompted with “How to write detailed queries for PubMed and PMC,” gpt-oss-120b invokes get_query_examples to provide database-agnostic query construction guidance, appropriately addressing both databases. gpt-oss-20b calls get_ pubmed_query_guide, focusing solely on PubMed-specific syntax despite the explicit mention of PMC in the user prompt, demonstrating reduced sensitivity to nuanced query requirements.

## 5 Discussion

This study demonstrates a proof-of-concept implementation of an LLM-based diagnostic decision support system that directly accesses EMR data and retrieves relevant case reports through a modular, MCP-based architecture. Our approach offers several key advantages for clinical deployment: reproducibility through open-source components and standardized interfaces, cost-effectiveness via on-premises LLM deployment, transparency through evidence linking that enables clinicians to trace system outputs to source data, and extensibility allowing integration of additional clinical resources such as guidelines and drug databases. By demonstrating this system on a diagnostically complex case, we show how structured EMR access and automated literature retrieval can support clinicians in generating differential diagnoses, particularly for rare or atypical presentations. We discuss how this approach aligns with clinical diagnostic workflows, examine model performance characteristics observed in our implementation, acknowledge important limitations, and outline considerations for broader clinical deployment.

### 5.1 Alignment with Clinical Diagnostic Process

Although initial diagnostic impressions may rapidly arise through System 1 reasoning triggered by salient patient cues, clinicians typically transition to a structured, analytic (System 2) process to refine and validate these early impressions before reaching a final diagnosis[57, 37]. This analytic process begins with a systematic review of vital signs, laboratory findings, and key information extracted from the EMR, through which clinicians generate an initial set of diagnostic hypotheses informed by their clinical knowledge and illness scripts. These hypotheses are verified or excluded by additional diagnostic tests, detailed history taking, or physical examinations. When the leading hypotheses are ruled out, the clinicians revisit the EMR, acquire further history, seek specialist consultation, and conduct reassessments to refine their diagnostic understanding. If diagnostic uncertainty persists even after reevaluation, clinicians review the case reports for such a rare or atypical diseases through resources such as PubMed, or UpToDate.

In this context, particularly when clinicians turn to external resources for rare or unfamiliar conditions, our DDx-Finder system can provide timely assistance at several points:

- Extract features from EMR that are masked by cognitive biases.
- Generate high-quality search queries for clinicians who have limited experience in reviewing case reports
- Quick and concise summaries of retrieved case reports

By supporting clinicians at these critical junctures, our system aligns well with their natural workflow and may help reduce diagnostic delays in complex or unfamiliar presentations. Notably, recent research demonstrates that diagnostic errors stem primarily from insufficient access to relevant knowledge rather than from cognitive biases[51, 42], highlighting the critical importance of tools that facilitate rapid access to pertinent case reports and evidence.

### 5.2 Comparison with Commercial Diagnostic Decision Support Systems

Established commercial platforms such as Isabel[36] and VisualDx[53] have demonstrated clinical utility through cloud-based subscription services, which require patient data transmission to external servers that institutions with strict data localization requirements must consider. VisualDx requires recurring fees from $19.99 to $38.99 monthly for individuals, with institutional costs substantially higher; Isabel similarly operates on subscription models. DDx-Finder eliminates these recurring costs and external data transmission by operating entirely within institutional infrastructure, requiring only initial GPU investment and routine maintenance. This modular architecture also permits institutions to adapt MCP components with AI-assisted development tools, facilitating iterative improvements that leverage advancing LLM capabilities while addressing local practice requirements. Additionally, while Isabel’s reference dataset updates monthly and VisualDx employs curated physician-edited content, DDx-Finder provides real-time PubMed access, particularly valuable for rare diseases and atypical presentations where curated databases may lag.

DDx-Finder may serve either as replacement or complementary system depending on institutional priorities. For institutions prioritizing data sovereignty, cost containment, or unbiased EMR interpretation, DDx-Finder provides a viable alternative to cloud-based platforms, potentially serving as the primary diagnostic support system across common and complex cases. Conversely, Isabel and VisualDx have maintained commercial operations for about twenty years with intermittent clinical validation studies supporting their use in specific contexts[55, 17, 14, 15, 16, 22]. Isabel demonstrates 96% accuracy within top 10 differentials[35] and VisualDx provides extensive image libraries showing disease variation[67], offering validated performance with immediate deployment. Institutions valuing specialty-specific features may implement hybrid deployment, leveraging VisualDx for dermatology imaging or Isabel for rapid differentials in time-constrained settings. DDx-Finder complements these systems by addressing diagnostically challenging cases requiring recent literature access, handling presentations inadequately covered by curated databases, or serving environments with data governance requirements or regulatory constraints that preclude cloud-based platforms.

### 5.3 Model Size and Clinical Reasoning Performance

Although systematic model comparison was not the primary focus of this work, our proof-of-concept case study revealed performance differences between the two models. For EMR-based clinical reasoning, gpt-oss-120b, demonstrating superior instruction adherence, produced structured outputs with context-appropriate terminology that accurately distinguished acute from chronic symptoms in clinical state extraction and employed quantitative evidence from laboratory and imaging data with broader consideration of cardiopulmonary and systemic etiologies in differential diagnosis generation. In contrast, gpt-oss-20b occasionally generated contextually inappropriate mappings and relied on qualitative imaging-centered reasoning focused on structural pathology, requiring more human refinement. For literature retrieval tasks, gpt-oss-120b constructed comprehensive search queries emphasizing acute presenting symptoms with proactive refinement guidance and maintained factual accuracy in abstract summarization and bibliographic link generation, whereas gpt-oss-20b focused on chronic disease complications and introduced factual errors with malformed citations, requiring careful attention during clinical reasoning. For literature retrieval tasks, gpt-oss120b constructed comprehensive search queries emphasizing acute presenting symptoms with proactive refinement guidance and maintained factual accuracy in abstract summarization and bibliographic link generation, whereas clinical reasoning with gpt-oss-20b outputs required greater attention due to these limitations.

As documented in Figure 4 of [52], gpt-oss-120b consistently outperformed gpt-oss-20b across HealthBench variants[52]. While both models showed performance differences on HealthBench (57.6% vs 42.5%) and HealthBench Consensus (90.0% vs 82.6%), the gap was most pronounced on HealthBench Hard—a challenging subset requiring complex clinical reasoning—where gpt-oss-120b scored 30.0% compared to gpt-oss-20b’s 10.8%, aligning with our observations of superior instruction adherence and factual accuracy in the larger model. However, Gu et al.[31] reported contrasting findings, showing that larger models demonstrated superior efficiency in diagnostic reasoning but not overwhelmingly so, with no substantial differences in factuality or completeness between model sizes (Supplementary Tables 4-5). This study did not include gpt-oss models, and performance varies considerably by model architecture and training. Notably, even advanced models achieved only 50% completeness in 1-turn reasoning, suggesting the benchmark may be too complex to reliably distinguish model-size effects. Given that our observations are based on a single case study while Gu et al. evaluated 1,453 cases, definitive conclusions about model size effects on clinical reasoning require further investigation.

### 5.4 Limitations

Despite the system’s utility, the limited token capacity of current LLMs poses challenges for extracting information from EMRs and literature. As lengthy, unstructured progress notes are recorded daily, summarizing them may exceed the model’s context limits, potentially omitting important abnormal values from laboratory results or losing nuanced symptom descriptions and critical assessments. Additionally, copy-forward documentation practices, where clinicians propagate content across sequential notes, introduce textual redundancy that unnecessarily consumes limited context windows. Similarly, summarizing long case report abstracts may fail to capture key details of rare diseases when retrieved text exceeds context limits. These limitations suggest that EMRs may need to evolve towards more structured, LLM-compatible formats to support secondary use, and that hierarchical summarization or prioritization strategies that retrieve clinically relevant information may be necessary.

This proof-of-concept study has important limitations regarding generalizability and evaluation. Our demonstration using a single complex case, while illustrative of the system’s capabilities, requires large-scale validation across diverse patient populations and clinical contexts. The manual evaluation approach lacks automated metrics for assessing whether appropriate case reports are retrieved given real-world EMR inputs, which would require goldstandard datasets that define expected relevant literature for specific clinical presentations. Additionally, our current implementation handles Korean EMR data, necessitating adaptation for English EMR systems and other languages. Longer timescales of EMR data may also pose challenges not evident in this single-encounter demonstration. However, the pipeline architecture is designed for generalizability: EMR formats can be adapted through configuration, MCP servers provide abstraction over data sources, and prompt engineering allows customization for different medical specialties.

Current EMR documentation practices present additional limitations for LLM-based diagnostic support. Progress notes, particularly their subjective sections, often lack detailed documentation of patient-physician interactions and patients’ subjective symptom experiences. While objective medical records enable differential diagnosis generation, richer documentation of patient narratives and clinical dialogue would significantly enhance our system’s diagnostic capabilities. Fortunately, this limitation may be addressed in the near future through emerging speech-to-text technologies that automatically capture and summarize patient-physician interactions into structured clinical notes[71, 19, 3], potentially providing the contextual richness needed for more comprehensive diagnostic reasoning.

Finally, it is essential to acknowledge that LLM-based diagnostic decision support systems, like their classical predecessors, require careful clinical interpretation. Early developers of computer-assisted diagnostic systems anticipated that these tools would provide convenient, inexpensive, and accurate second opinions [12, 28]. However, experience has shown that such systems generate numerous false positives and potentially miss critical diagnoses (false negatives), necessitating expert clinical judgment to interpret results appropriately. LLM-based DDx support systems face similar challenges[12, 28], and clinical expertise remains indispensable for validating system outputs, identifying spurious suggestions, and recognizing overlooked diagnoses. Our system should therefore be viewed as a supportive tool that augments rather than replaces clinical reasoning.

### 5.5 Considerations for Clinical Deployment

Encouragingly, our proposed system architecture is immediately deployable for beta testing within our institution once database access permissions are granted, as the modular MCP-based design requires no substantial modifications to existing clinical infrastructure. However, broader clinical deployment across diverse healthcare settings will require addressing several engineering and operational challenges. Hospitals will need efficient pipelines to extract useful records from EMR database efficiently, while preserving temporal fidelity and data integrity. Institutions might also determine which scenarios are appropriate and which documentations or note types should be optimized for downstream usage. In addition, deploying LLMs in clinical environments requires substantial computational resources, particularly when processing large amount of EMR data or maintaining long-context windows to handle. Scalability, cost, security, and governance frameworks must be established to ensure safe integration into clinical workflows. These considerations underscore the needs for a coordinated infrastructure before LLM-based clinical assistants can be reliably adopted in practice.

## 6 Future Work

We are planning:

1. **Expanded single-center validation**: 100+ cases across multiple specialties at Asan Medical Center
2. **Quantitative evaluation**: Precision, recall, and diagnostic accuracy metrics against gold standards
3. **Baseline comparisons**: Performance comparison with GPT-5, Claude, and rule-based systems
4. **User studies**: Clinician feedback on system usability and clinical utility
5. **Real-time deployment**: Integration into clinical workflow for prospective evaluation

## 7 Conclusion

DDx-Finder demonstrates the feasibility of combining on-premises LLMs with MCP servers to provide clinicians with automated EMR analysis and literature retrieval for differential diagnosis support. Our modular architecture offers a reproducible, transparent, and cost-effective foundation that augments clinical reasoning while preserving clinical judgment. The system aligns with analytic diagnostic workflows and is immediately deployable for institutional testing. While this proof-of-concept validates the technical approach on a complex case, large-scale clinical evaluation is needed to assess real-world impact. We release our code and documentation to enable community development of human-in-the-loop differential diagnosis systems.

## Data Availability

All data produced in the present study are available upon reasonable request to the authors

## Ethics Statement

This study evaluated a technological framework (MCP-based summarization system) using a fully de-identified single clinical case for technical validation. As the study utilized strictly anonymized retrospective data without secondary patient risk, it was determined to be exempt from formal review by the Institutional Review Board (IRB) of Asan Medical Center, Seoul, Korea (IRB Exemption No. 2026-1156) in accordance with institutional guidelines and national regulations.

## Informed Consent Statement

Patient consent was waived by the IRB due to the retrospective nature of the study using fully anonymized data with no potential risk to patient privacy.

## Code Availability

Source code and documentation are available at: https://github.com/loopback-kr/DDx-Finder

### A EMR Processing Algorithms

This appendix provides detailed pseudocode for the core EMR processing algorithms used in DDx-Finder’s summarize_ medical_records function. The algorithms implement patient-independent extraction strategies that rely on visual markers and structural section headers rather than disease-specific medical knowledge.

#### Algorithm 1 Intelligent Medical Record Summarization

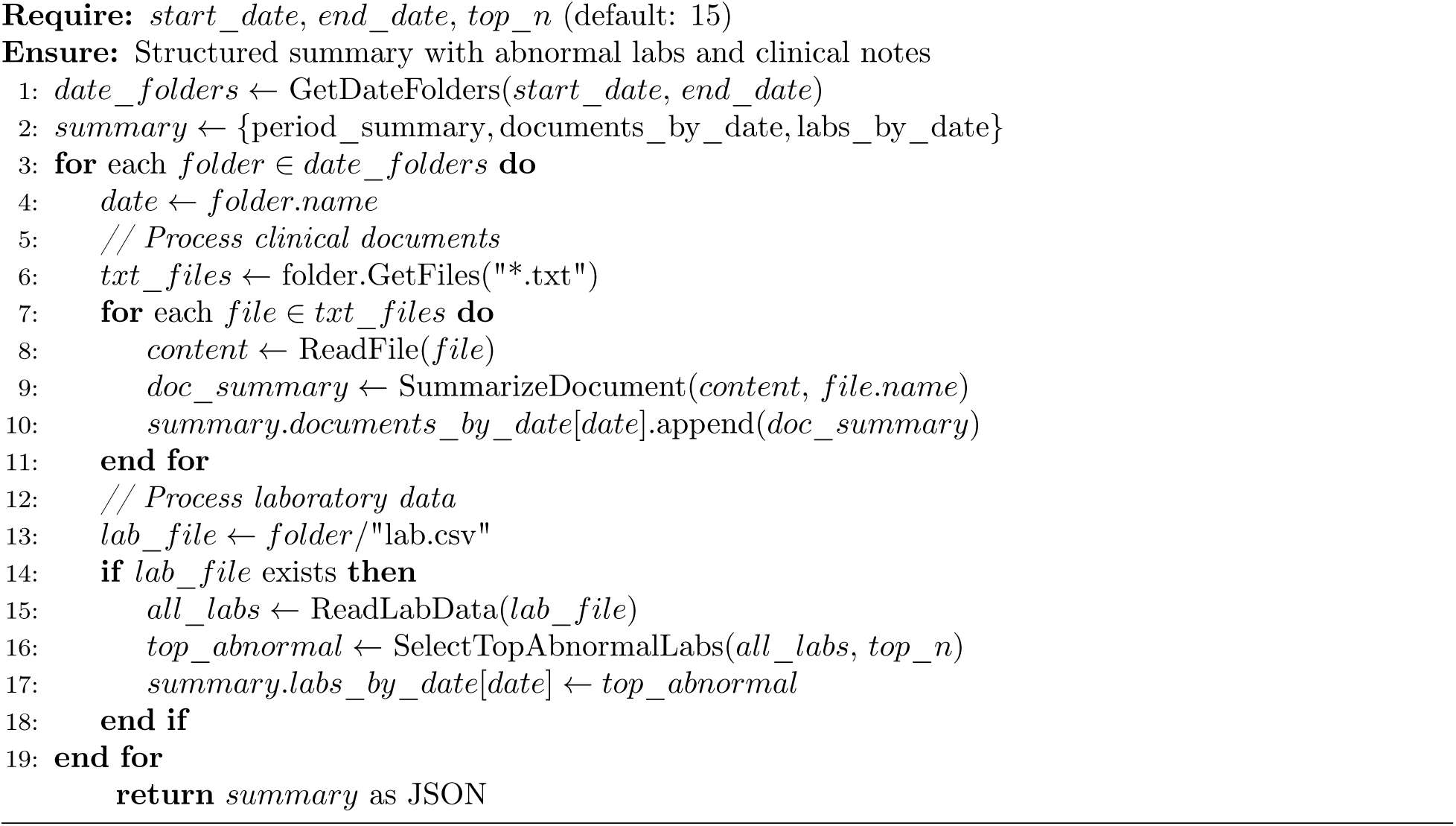

#### Algorithm 2 Select Top Abnormal Laboratory Results

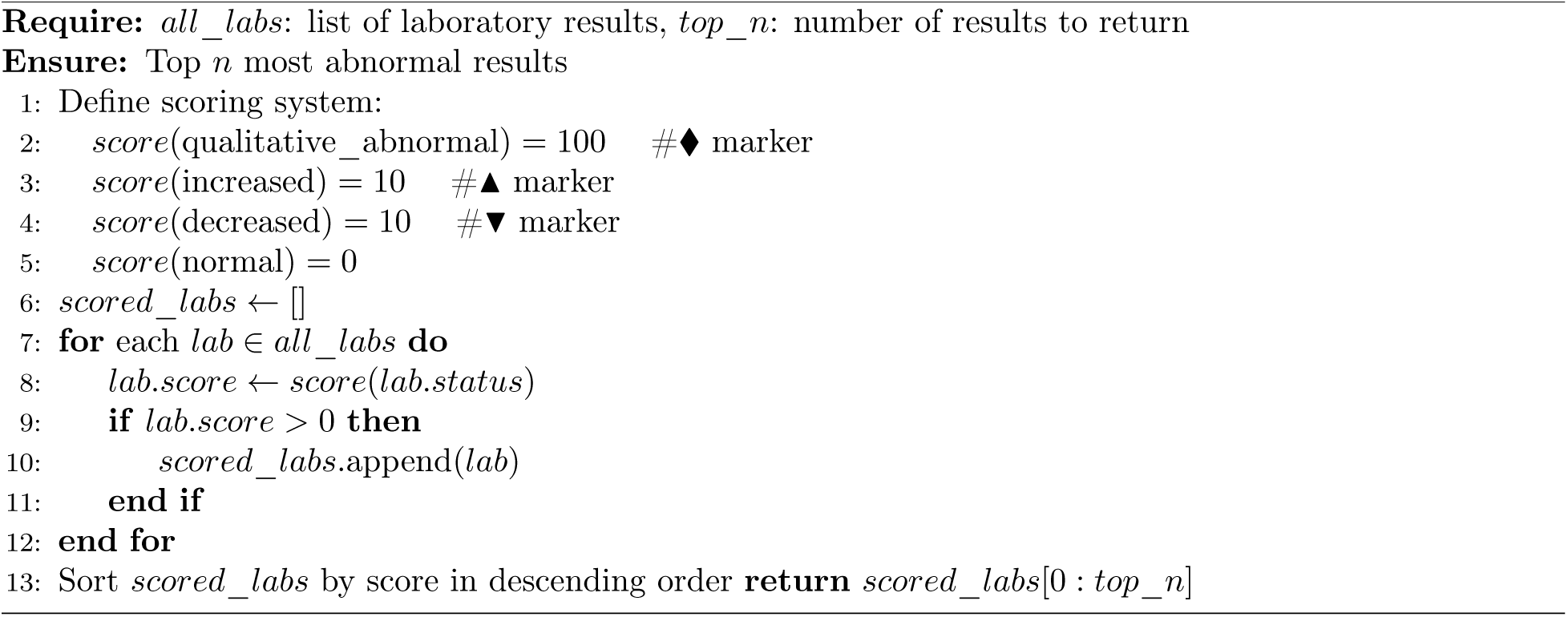

#### Algorithm 3 Summarize Clinical Document Structure

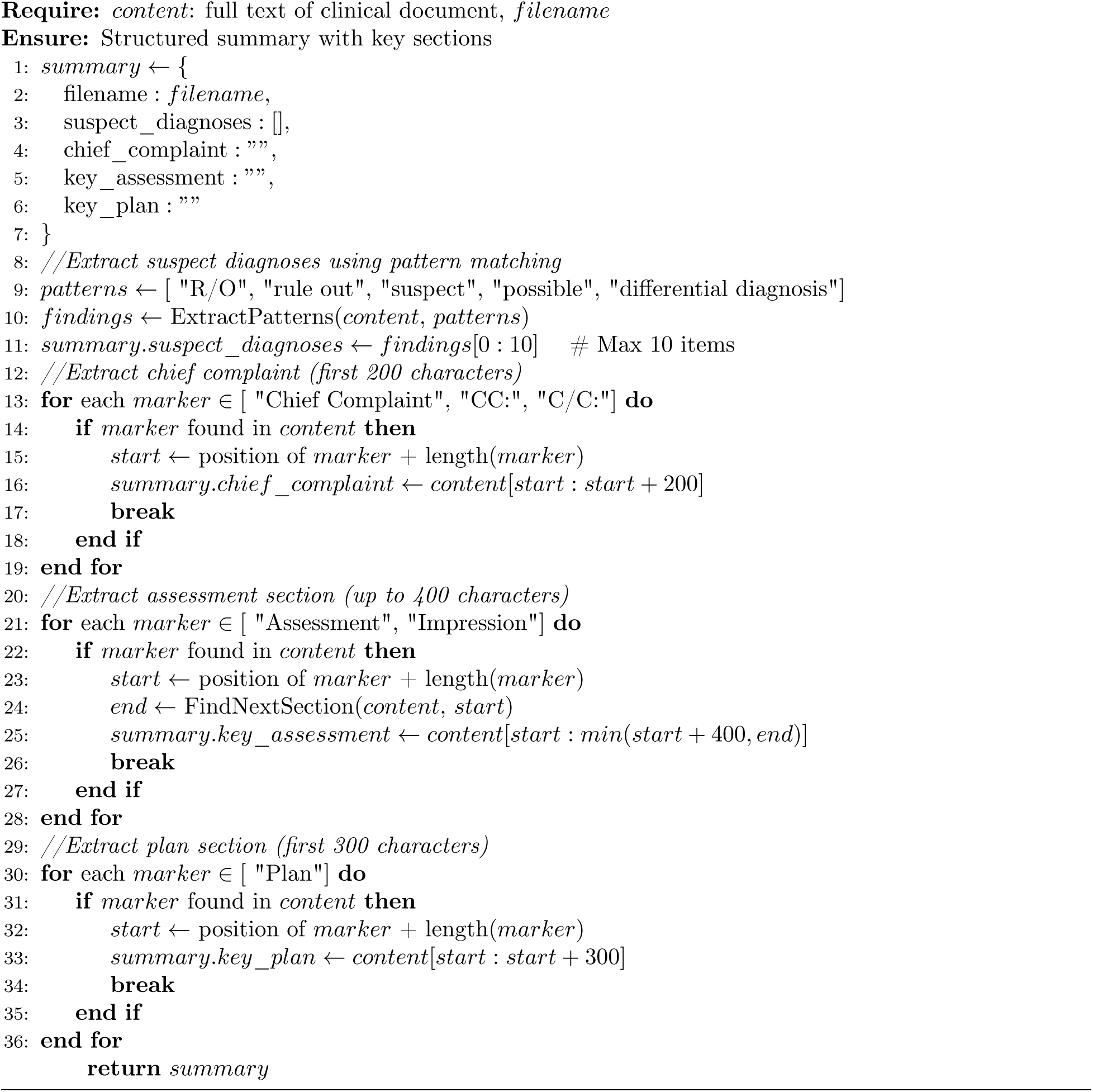

**Algorithm Characteristics** The algorithms implement the following design principles:

- **Patient-independent**: No patient-specific hardcoding; applicable to any clinical case
- **Structure-based extraction**: Relies on visual markers (♦, ▴, ▾) and section headers rather than specific medical knowledge
- **Token-efficient**: Returns only top abnormal findings, reducing token consumption by approximately 20% while preserving clinical significance
- **Relative importance**: Prioritizes findings within the patient’s own data distribution rather than absolute reference ranges

### B Literature Search Algorithms

This appendix provides detailed pseudocode for the core literature search algorithms used in DDx-Finder’s search_literature function. The algorithms implement multi-database querying with query transparency and token-efficient result presentation.

#### Algorithm 4 Multi-Database Literature Search

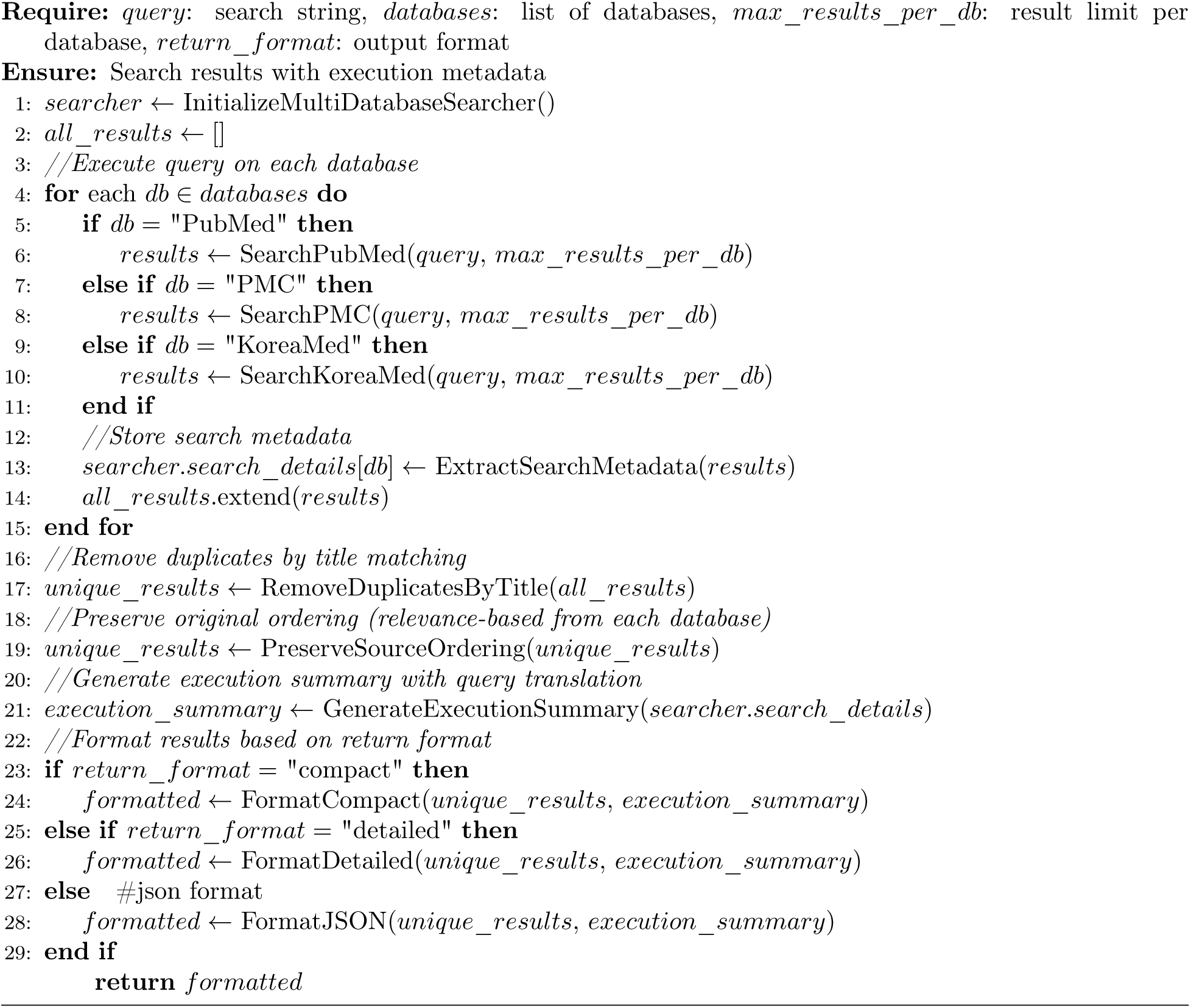

#### Algorithm 5 Search Single Database (PubMed/PMC)

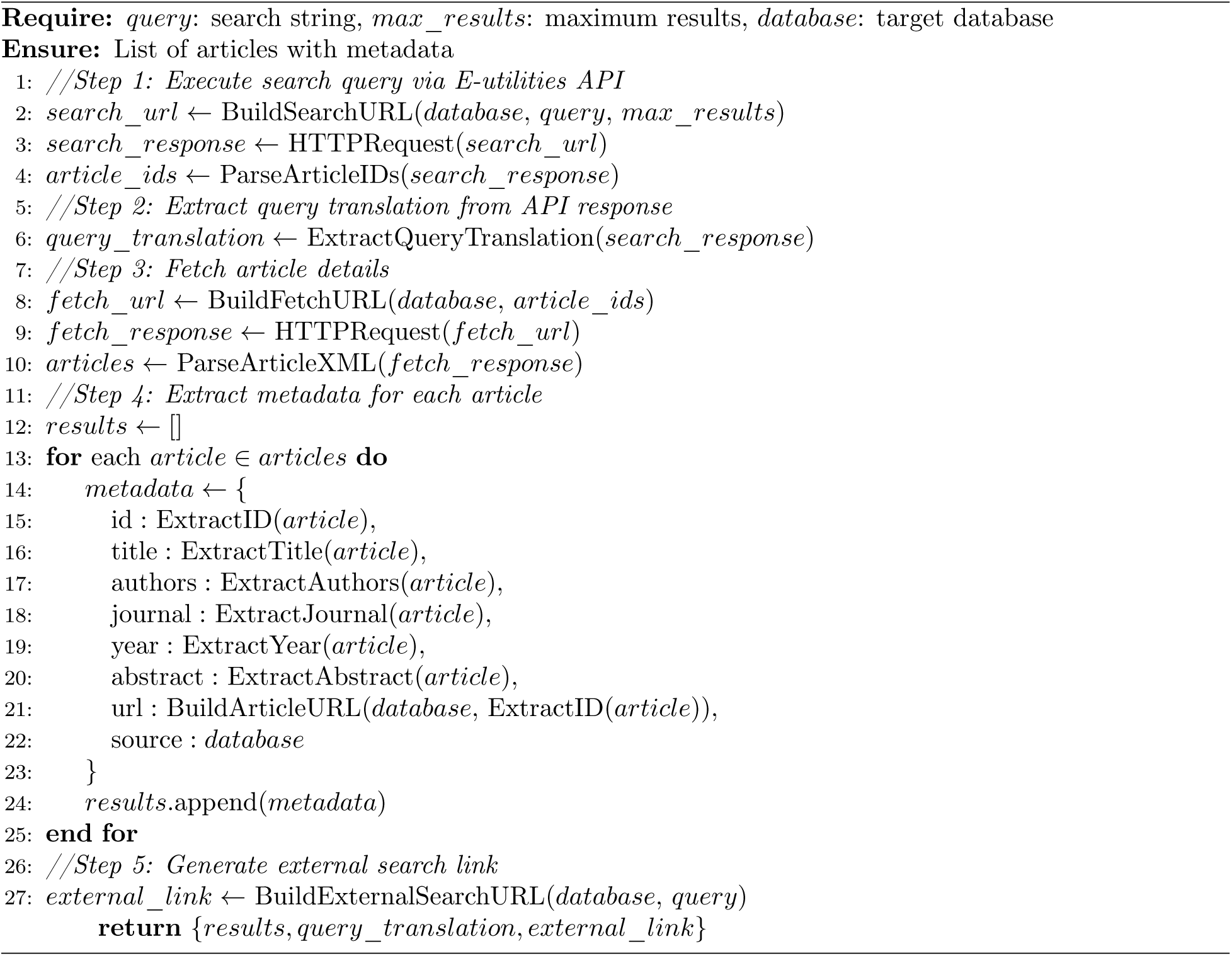

#### Algorithm 6 Token-Efficient Result Formatting

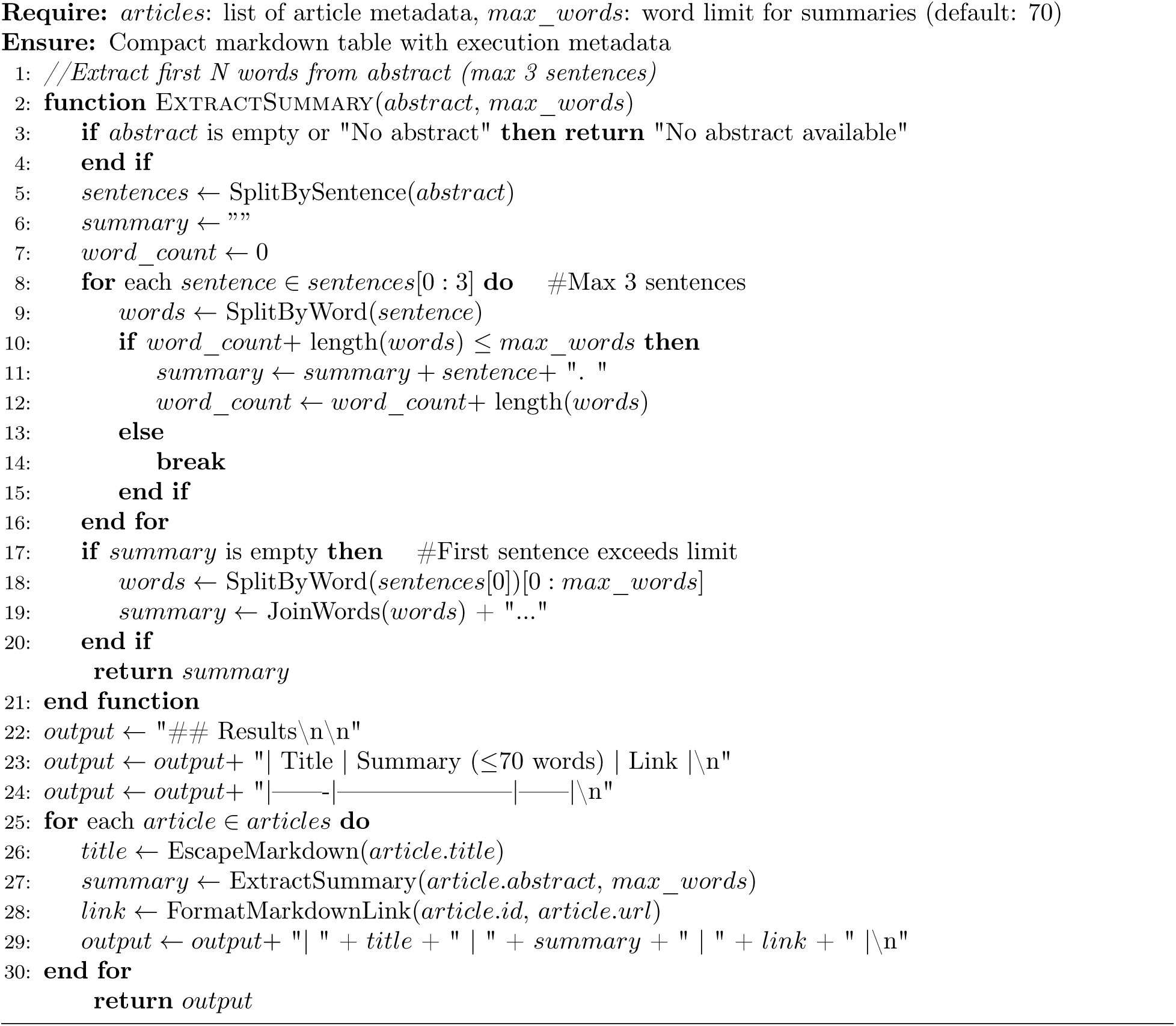

#### Algorithm 7 Remove Duplicate Articles by Title

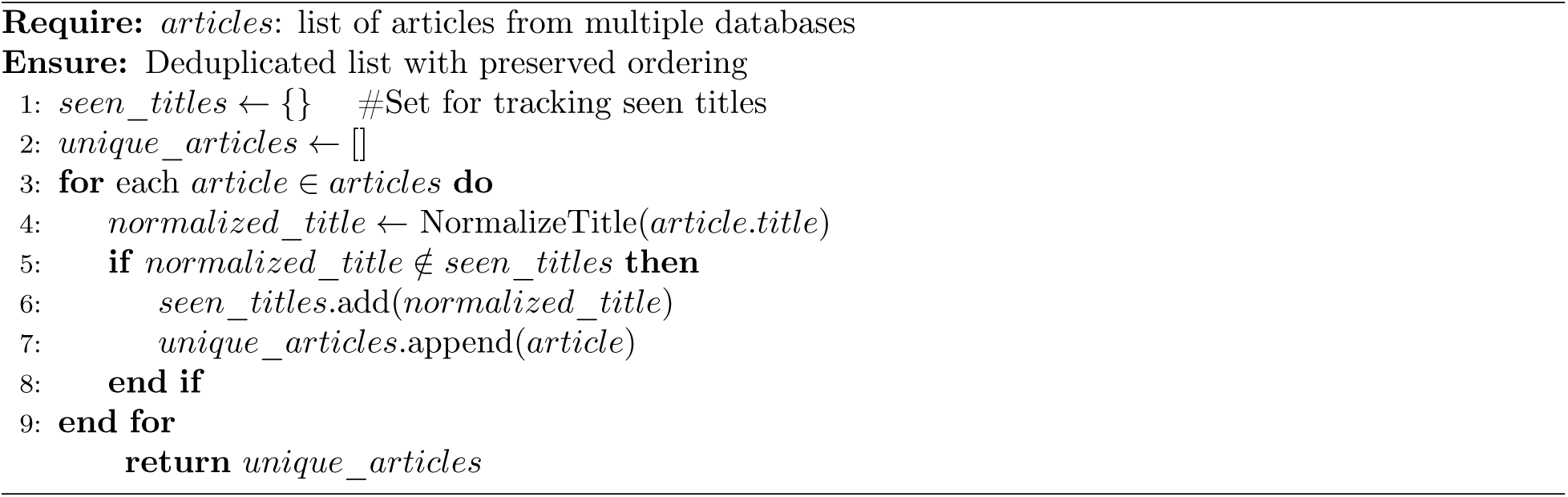

#### Algorithm 8 Generate Execution Summary with Query Translation

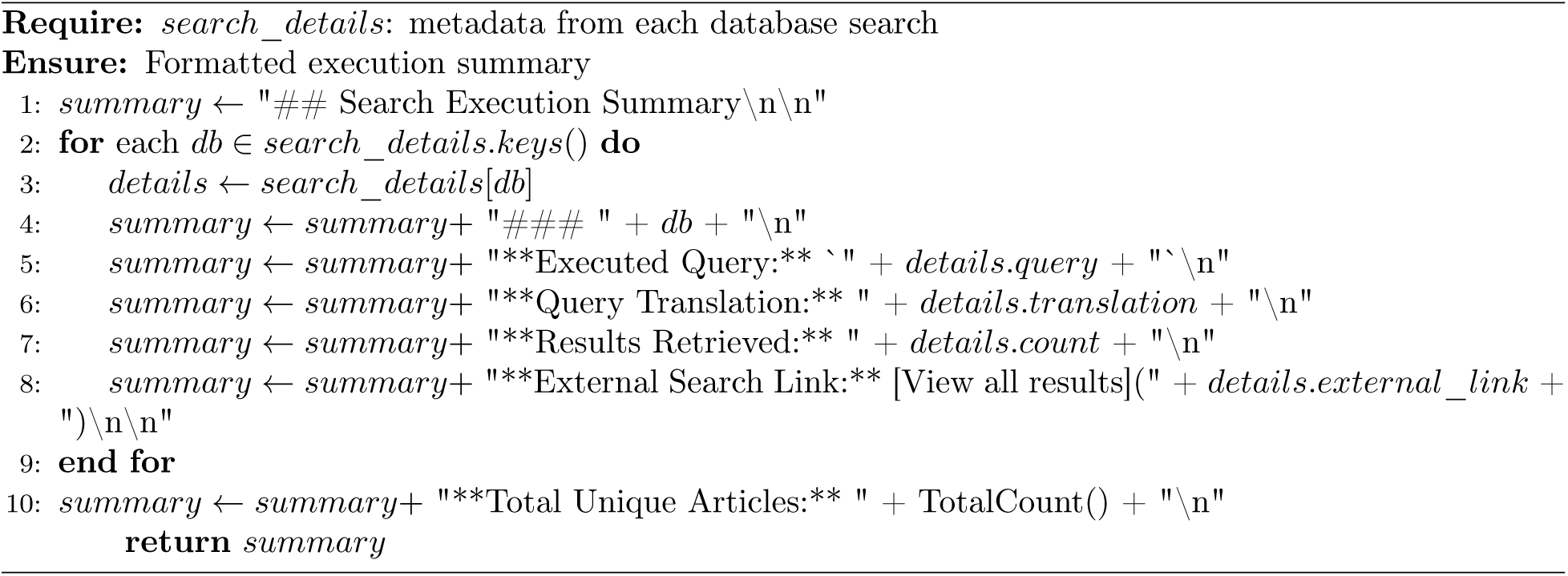

**Algorithm Characteristics** The literature search algorithms implement the following design principles:

- **Query transparency**: The exact query string is sent to each database without modification or simplification. Query translation from each search engine shows how the query was interpreted, enabling iterative refinement.
- **Multi-database execution**: Queries are executed in parallel across PubMed (abstracts), PMC (full-text), and optionally KoreaMed (Korean literature). Each database’s relevance-based ordering is preserved.
- **Token efficiency**: Compact format extracts only the first 70 words (maximum 3 sentences) from abstracts, reducing token consumption by 60-80% compared to displaying complete abstracts while preserving sufficient information for relevance assessment.
- **Configurable output formats**: Three formats accommodate different use cases:

- *Compact* (default): Execution metadata + article table with truncated summaries
- *Detailed* : Execution metadata + full abstracts for each article
- *JSON* : Structured data with separate execution summary field
- **Duplicate removal**: Articles appearing in multiple databases are identified by normalized title matching and deduplicated while preserving the first occurrence’s ordering.
- **External search links**: Direct URLs to database search pages enable exploration of results beyond token limits, supporting comprehensive literature review.
- **Iterative refinement support**: Query translation metadata reveals automatic term mapping and field tag resolution, enabling informed query adjustment when initial results are insufficient or off-target.

#### B.1 Token Efficiency Analysis

The compact format achieves significant token reduction through mechanical abstract truncation:

**Table B.1:**
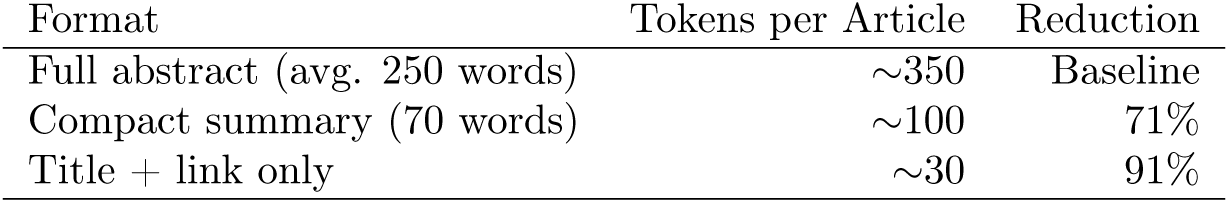
Token Consumption Comparison.

The 70-word threshold balances token efficiency with clinical utility:

- Sufficient context for relevance assessment
- Preserves key clinical presentation details
- Enables rapid screening of 20+ articles within typical context windows
- LLM can request full abstract via external link if needed

The configurable max_words parameter allows adjustment based on available context window and result set size, with 70 words empirically determined as optimal for most clinical queries.

### C Case Study Interaction Transcript

This appendix provides the complete interaction transcript between the clinician and DDx-Finder for the case study presented in Section 4. Each turn includes the user prompt and the system response with supporting data.

#### C.1 Initial Clinical State Extraction

##### C.1.1 User Prompt

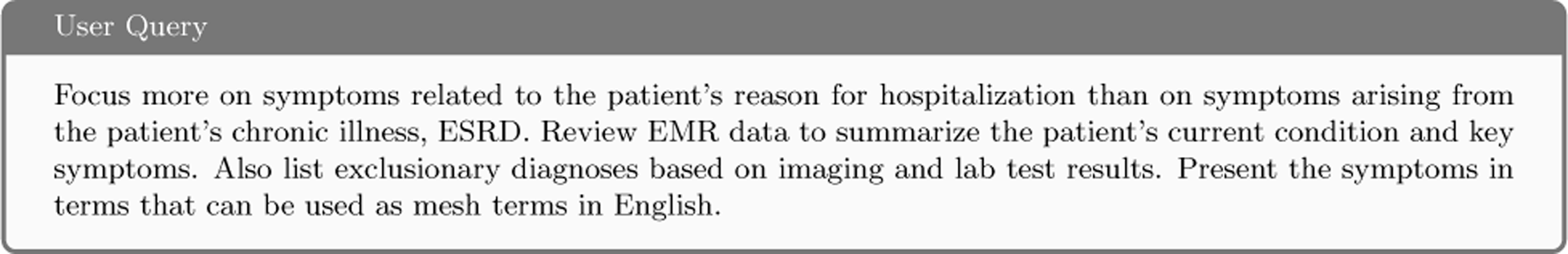

##### C.1.2 gpt-oss-120b Response

**Current Condition (Focus on Acute Reason for Admission)** The system extracted the following clinical state organized by domain:

**Table C.1:** Current Clinical Condition Extracted from EMR.

| Domain | Key Findings (derived from EMR) | MeSH-compatible terms |
| --- | --- | --- |
| Gastro-intestinal | Worsening liver-function tests (AST/ALT $\uparrow \approx 80$–112 IU/L)<br>Frequent watery diarrhea noted in admission note<br>Superficial and atrophic gastritis on EGD<br>Nonspecific ileitis on colonoscopy<br>Bowel wall edema on CT (no obstruction) | Diarrhea; Gastritis, Atrophic; Gastritis, Superficial; Ileitis, Nonspecific; Bowel Edema |
| Musculoskeletal / pain | Left-thigh (femoral) pain causing sleep disturbance (admission note)<br>Hip pain (progress notes) with no bony abnormality on plain OS<br>Rhabdomyolysis suspected (CK $\approx 5,300$–1,500 IU/L, Myoglobin $\approx 4,400$–8,600 ng/mL) | Thigh Pain; Hip Pain; Rhabdomyolysis; Myalgia; Sleep Disturbance |
| Constitutional / neuro-logic | Generalized weakness/fatigue (“Asthenia”) starting 6 months before admission<br>Stress-related concentration difficulty | Fatigue; Weakness; Stress, Psychological |
| Metabolic / electrolyte | Hyponatremia (Na $\approx 124$–130 mmol/L)<br>Hyperphosphatemia (P $\approx 5.0$–7.2 mg/dL)<br>Very low albumin ($\approx 1.8$ g/dL)<br>Elevated PTH (875 pg/mL) | Hyponatremia; Hyperphosphatemia; Hypoalbuminemia; Hyperparathyroidism |
| Renal / urinary | End-stage renal disease on CAPD (baseline) – not the primary admission problem<br>Complicated renal cysts/Bosniak IV cysts on CT; acquired cystic kidney disease listed repeatedly | Renal Cysts, Complicated; Acquired Cystic Kidney Disease |
| Other | Inguinal hernia scheduled for repair<br>GERD symptoms noted (“GERD complaint”) | Inguinal Hernia; Gastroesophageal Reflux |

**Exclusionary Diagnoses (Based on Imaging, Labs, and Clinical Data)** The system identified conditions that could be excluded based on available evidence:

**Table C.2:** Exclusionary Diagnoses from EMR Analysis.

| Condition | Reason for Exclusion (Evidence) |
| --- | --- |
| Acute myocardial infarction / coronary artery disease | CK-MB only mildly ↑ (71 ng/mL); normal transthoracic echo (no wall-motion abnormality) and no ST-segment changes reported on EKG. |
| Pulmonary embolism | D-dimer modestly ↑ (1.73 $\mu\text{g/mL}$ FEU) but no CT-angiography or V/Q study; no clinical signs of right-heart strain on echo. |
| Acute bowel obstruction | CT shows bowel edema but no dilated loops, transition point, or obstruction; colonoscopy and EGD are unremarkable except for gastritis/ileitis. |
| Severe infectious sepsis | No positive blood cultures, viral hepatitis serologies negative (Anti-HCV non-reactive, HCV RNA not detected), syphilis non-reactive; inflammatory markers not markedly elevated. |
| Acute hepatitis B/C | Anti-HCV negative, HCV RNA not detected; no hepatitis B data but liver enzyme elevation is modest and could be drug- or uremia-related. |
| Cardiac tamponade / pericardial effusion | Normal echo with preserved chamber dimensions; no pericardial fluid reported. |
| Pancreatitis | No amylase/lipase reported, imaging does not show pancreatic swelling or fluid. |
| Severe electrolyte derangement requiring emergent correction | Hyponatremia present but mild-moderate ($\text{Na} \approx 124\text{--}130$ mmol/L) and stable; no seizures or coma documented. |

**Summary** The patient was admitted primarily for worsening liver-function tests, frequent diarrhea, and left-thigh pain that interfered with sleep—symptoms distinct from the chronic end-stage renal disease (ESRD) background. Laboratory work shows a pattern of hepatic injury, marked muscle breakdown (rhabdomyolysis), hyponatremia, hyperphosphatemia, and severe hypoalbuminemia, while imaging rules out acute cardiac, pulmonary, or obstructive abdominal pathology. The current clinical picture centers on gastrointestinal irritation (diarrhea, gastritis, ileitis), musculoskeletal pain (thigh/hip pain, rhabdomyolysis), and metabolic derangements, all of which can be indexed with the MeSH terms listed above.

*Note: All data referenced are taken from the EMR summary provided by the summarize_ medical_ records tool*.

##### C.1.3 gpt-oss-20b Response

**Current Condition & Key Symptoms (Focus on Reason for Admission)** The system extracted the following clinical symptoms from the EMR data:

**Table C.3:**
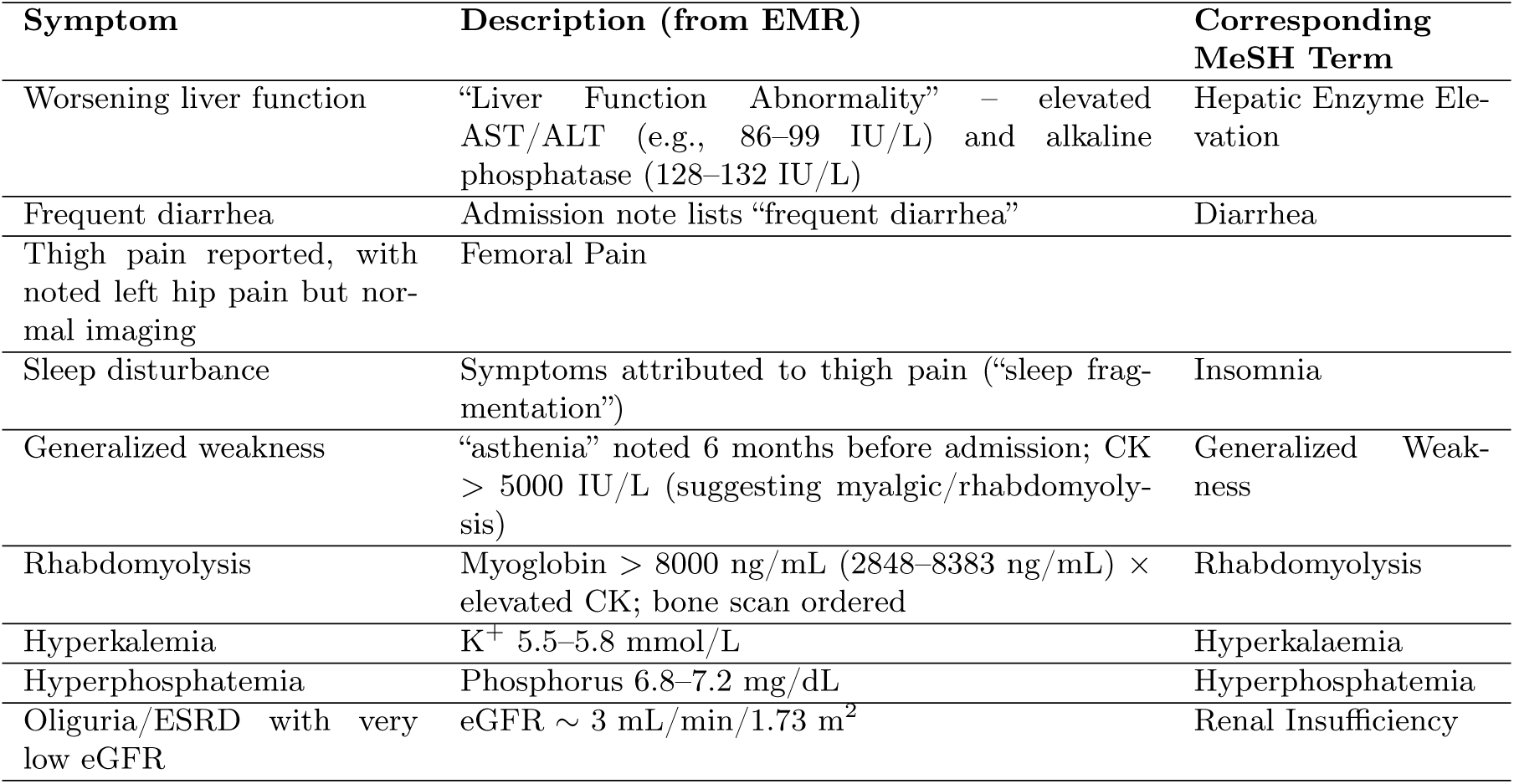
Clinical State Extraction by gpt-oss-20b.

**Exclusionary Diagnoses (Imaging & Laboratory Findings)** The system identified conditions that could be excluded based on available evidence:

**Table C.4:** Exclusionary Diagnoses by gpt-oss-20b.

| Exclusionary Finding | Rationale |
| --- | --- |
| Acute pyelonephritis / urinary tract infection | CT shows cystic changes but no abscess or hydronephrosis; urine cultures not positive in data. |
| Acute appendicitis | No evidence of appendicular inflammation on CT; no right lower abdominal pain reported. |
| Acute mesenteric ischemia | CT shows low-density wall thickening (“proctocolitis”), no vascular occlusion; bowel sounds present. |
| Severe hepatic failure | AST/ALT modestly elevated; bilirubin not reported as high; clinical picture dominated by renal failure. |
| Skeletal fractures | Chest/abdominal CT and bone scan show no fractures; bone scan indicates acute rhabdomyolysis, not bone injury. |
| Vascular aneurysm | CT of abdomen does not reveal any aneurysmal dilation; only cystic kidney lesions. |

**Summary** These exclusions are supported by the imaging (CT, bone scan, echocardiogram) and laboratory data displayed for the 8-day period. The primary acute issues are rhabdomyolysis with associated hyperkalaemia, hyperphosphatemia, and hepatic enzyme elevation, underpinned by the patient’s chronic end-stage renal disease.

*Note: All data referenced are taken from the EMR summary provided by the summarize_ medical_ records tool*.

#### C.2 Turn 2: Literature Search Query Generation

##### C.2.1 User Prompt

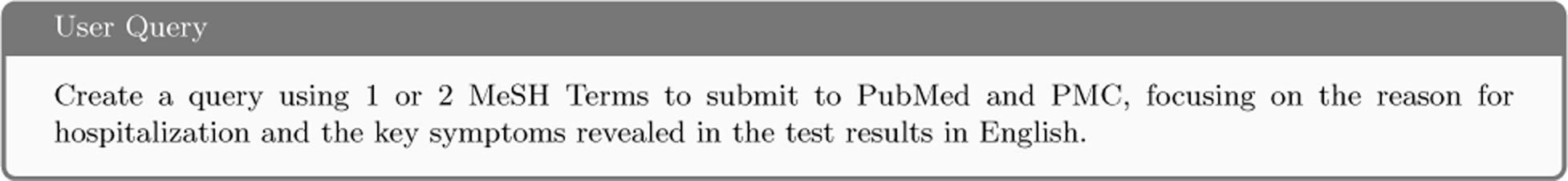

##### C.2.2 gpt-oss-120b Response

**PubMed / PMC Search – Two-MeSH-Term Query** Below is a ready-to-copy search string that uses exactly two MeSH headings and captures the acute problem that brought the patient to the hospital (muscle breakdown + gastro-intestinal disturbance) together with the most prominent laboratory-derived symptom (hyponatremia). You can paste the whole line into the PubMed or PubMed Central search box; the same syntax works on both sites.

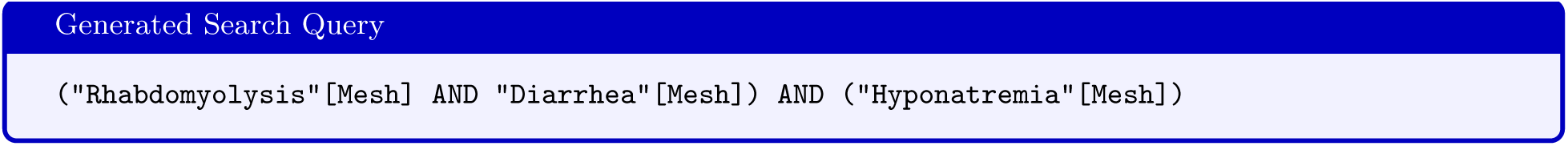

**How the Query Matches the Case** The generated query incorporates three key MeSH terms that directly correspond to the patient’s clinical presentation:

- **Rhabdomyolysis** – CK *≈* 5,300–1,500 IU/L, myoglobin *≈* 4,400–8,600 ng/mL *→* acute muscle injury that caused severe thigh/hip pain and sleep disturbance.
- **Diarrhea** – Frequent watery stools recorded in the chart (Mar 8–Mar 15) and ileitis on colonoscopy *→* a chief gastrointestinal complaint.
- **Hyponatremia** – Persistently low serum Na^+^ (*≈* 124–130 mmol/L) *→* a metabolic abnormality that often cooccurs with both rhabdomyolysis and severe diarrhea and contributed to the patient’s weakness.

The combination forces the search engine to return articles that discuss all three clinical entities together, which mirrors the patient’s acute presentation and therefore narrows the literature to the most pertinent evidence (e.g., case series of rhabdomyolysis precipitated by fluid-loss diarrhea, electrolyte disturbances, or both).

##### C.2.3 gpt-oss-20b Response

###### PubMed / PMC Search – Two-MeSH-Term Query

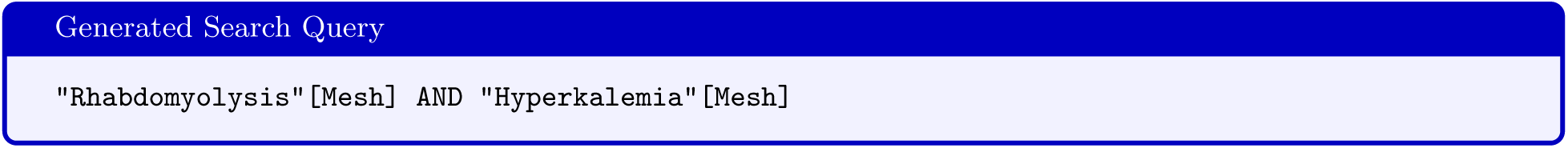

###### Explanation

- **Rhabdomyolysis** – captures the acute muscle-cell injury that is the patient’s primary cause of hospitalization, as demonstrated by the markedly elevated creatine kinase and myoglobin levels.
- **Hyperkalemia** – pulls in studies where the neurological, cardiac, and renal sequelae of high serum potassium (the patient’s life-threatening electrolyte disturbance) are examined. This two-term query will retrieve literature that specifically addresses the acute, life-threatening pathophysiology (rhabdomyolysis) that underlies the patient’s current hospitalization and the key laboratory manifestation (hyperkalemia).

This two-term query will retrieve literature that specifically addresses the acute, life-threatening pathophysiology (rhabdomyolysis) that underlies the patient’s current hospitalization and the key laboratory manifestation (hyperkalemia).

#### C.3 Literature Search for Case Reports

##### C.3.1 User Prompt

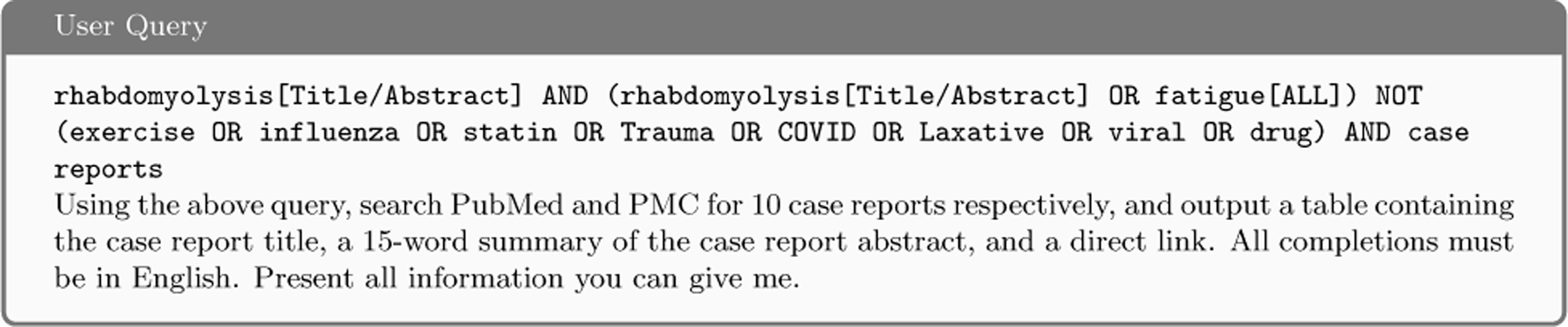

##### C.3.2 gpt-oss-120b Response

The system executed the literature search query across PubMed and PMC databases, retrieving 10 case reports from each source.

###### PubMed – 10 Case Reports

**Table C.5:** Case Reports Retrieved from PubMed.

| # | Title | 15-word Summary | Direct Link |
| --- | --- | --- | --- |
| 1 | We present a case report of rhabdomyolysis after intense physical activity... | Intense exercise caused CK rise, muscle pain, and marked AST/ALT elevation suggesting muscle-derived liver enzyme increase. | PMID:36896606 |
| 2 | Muscle Channelopathies and Rhabdomyolysis | Review of rare skeletal-muscle channel disorders that precipitate episodic rhabdomyolysis and diagnostic strategies. | PMID:41037179 |
| 3 | A Rare Case of Rhabdomyolysis | Describes localized myalgia, CK elevation, myoglobinuria, and potential progression to acute kidney injury. | PMID:36540529 |
| 4 | Rhabdomyolysis | Brief report without abstract; name-only entry indicating a documented rhabdomyolysis case. | PMID:3522983 |
| 5 | Olanzapine induced hyponatremia and rhabdomyolysis | Sudden hyponatremia from olanzapine triggered severe rhabdomyolysis, requiring ICU care. | PMID:36865653 |
| 6 | Probable chlorthalidone-induced hypokalemic rhabdomyolysis | Chlorthalidone dose increase caused profound hypokalemia and CK surge, confirming drug-related rhabdomyolysis. | PMID:39278350 |
| 7 | Meropenem Induced Hypokalemia | Two patients developed meropenem-related hypokalemia; case report focuses on electrolyte disturbance, not rhabdo. | PMID:28969183 |
| 8 | Hypothyroid-Induced Rhabdomyolysis: A Case Report | Severe hypothyroidism presented with generalized weakness, CK rise, and rhabdomyolysis, resolved after thyroid therapy. | PMID:39262557 |
| 9 | Schizoaffective Disorder and Concurrent Rhabdomyolysis | Psychiatric decompensation plus heat exposure led to rhabdomyolysis and acute renal injury. | PMID:34976507 |
| 10 | Severe Rhabdomyolysis in Glucose-6-Phosphate Dehydrogenase Deficiency | G6PD deficiency associated with rare rhabdomyolysis and myoglobinuria; case highlights genetic susceptibility. | PMID:32448501 |

###### PMC – 10 Case Reports

**Table C.6:**
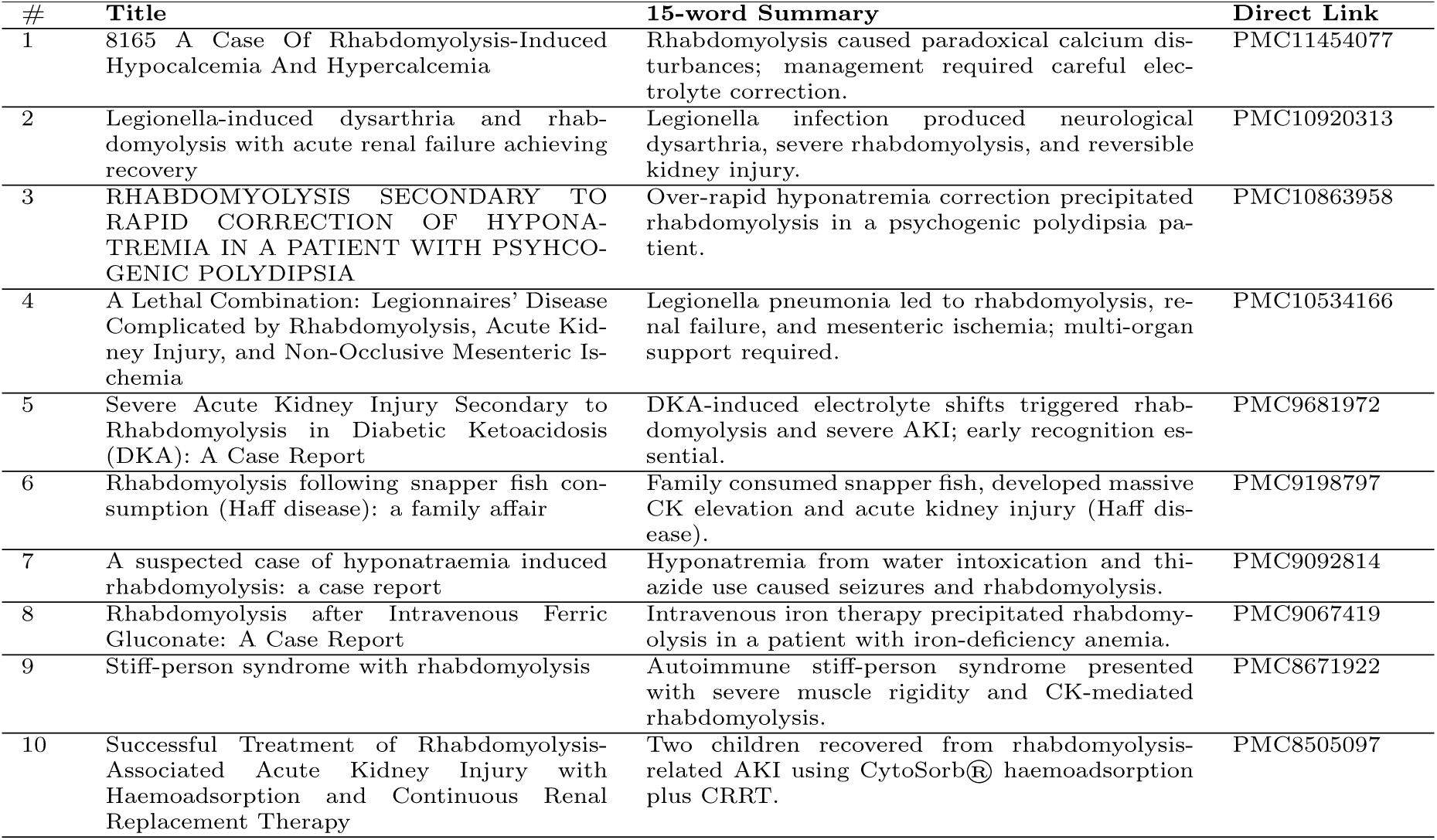
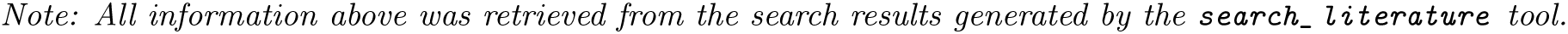
Case Reports Retrieved from PMC.

| # | Title | 15-word Summary | Direct Link |
| --- | --- | --- | --- |
| 1 | 8165 A Case Of Rhabdomyolysis-Induced Hypocalcemia And Hypercalcemia | Rhabdomyolysis caused paradoxical calcium disturbances; management required careful electrolyte correction. | PMC11454077 |
| 2 | Legionella-induced dysarthria and rhabdomyolysis with acute renal failure achieving recovery | Legionella infection produced neurological dysarthria, severe rhabdomyolysis, and reversible kidney injury. | PMC10920313 |
| 3 | RHABDOMYOLYSIS SECONDARY TO RAPID CORRECTION OF HYPONATREMIA IN A PATIENT WITH PSYCHOGENIC POLYDIPSIA | Over-rapid hyponatremia correction precipitated rhabdomyolysis in a psychogenic polydipsia patient. | PMC10863958 |
| 4 | A Lethal Combination: Legionnaires' Disease Complicated by Rhabdomyolysis, Acute Kidney Injury, and Non-Occlusive Mesenteric Ischemia | Legionella pneumonia led to rhabdomyolysis, renal failure, and mesenteric ischemia; multi-organ support required. | PMC10534166 |
| 5 | Severe Acute Kidney Injury Secondary to Rhabdomyolysis in Diabetic Ketoacidosis (DKA): A Case Report | DKA-induced electrolyte shifts triggered rhabdomyolysis and severe AKI; early recognition essential. | PMC9681972 |
| 6 | Rhabdomyolysis following snapper fish consumption (Haff disease): a family affair | Family consumed snapper fish, developed massive CK elevation and acute kidney injury (Haff disease). | PMC9198797 |
| 7 | A suspected case of hyponatraemia induced rhabdomyolysis: a case report | Hyponatremia from water intoxication and thiazide use caused seizures and rhabdomyolysis. | PMC9092814 |
| 8 | Rhabdomyolysis after Intravenous Ferric Gluconate: A Case Report | Intravenous iron therapy precipitated rhabdomyolysis in a patient with iron-deficiency anemia. | PMC9067419 |
| 9 | Stiff-person syndrome with rhabdomyolysis | Autoimmune stiff-person syndrome presented with severe muscle rigidity and CK-mediated rhabdomyolysis. | PMC8671922 |
| 10 | Successful Treatment of Rhabdomyolysis-Associated Acute Kidney Injury with Haemoadsorption and Continuous Renal Replacement Therapy | Two children recovered from rhabdomyolysis-related AKI using CytoSorb® haemoadsorption plus CRRT. | PMC8505097 |

*Note: All information above was retrieved from the search results generated by the `search_literature` tool.*

##### C.3.3 gpt-oss-20b Response

The system executed the literature search query across PubMed and PMC databases, retrieving 10 case reports from each source.

###### PubMed – 10 Case Reports

**Table C.7:** Case Reports Retrieved from PubMed by gpt-oss-20b.

| # | Title | 15-word Summary | Direct Link |
| --- | --- | --- | --- |
| 1 | We present a case report of rhabdomyolysis after intense physical activity... | Patient developed rhabdomyolysis after intense exercise; CK, AST, ALT rose, suspected liver injury requiring hospitalization. | PMID:36896606 |
| 2 | Muscle Channelopathies and Rhabdomyolysis | Article reviews channelopathies causing episodic rhabdomyolysis, discusses genetics, diagnosis, and management for patients in clinical. | PMID:41037179 |
| 3 | A Rare Case of Rhabdomyolysis | Case describes generalized myalgia, weakness, high CK, myoglobinuria, and risk of kidney injury in patient. | PMID:36540529 |
| 4 | Rhabdomyolysis | Brief report with no abstract, describes rhabdomyolysis presentation, diagnostic imaging, and treatment approach for patients. | PMID:3522983 |
| 5 | Olanzapine induced hyponatremia and rhabdomyolysis | Case of olanzapine therapy leading to hyponatremia and severe rhabdomyolysis, necessitating ICU care for treatment. | PMID:36865653 |
| 6 | Probable chlorthalidone-induced hypokalemic rhabdomyolysis | Patient on chlorthalidone developed hypokalemia and rhabdomyolysis; thorough workup excluded other causes through comprehensive evaluation. | PMID:39278350 |
| 7 | Meropenem Induced Hypokalemia | Meropenem use caused hypokalemia; not directly rhabdomyolysis but can worsen muscle injury in patients treatment. | PMID:28969183 |
| 8 | Hypothyroid-Induced Rhabdomyolysis: A Case Report | Hypothyroid patient presented with weakness, myalgia, and rhabdomyolysis; thyroid replacement reversed symptoms in elderly patient. | PMID:39262557 |
| 9 | Schizoaffective Disorder and Concurrent Rhabdomyolysis | Psychiatric exacerbation accompanied by rhabdomyolysis and renal failure after heat exposure in patient requiring ICU. | PMID:34976507 |
| 10 | 8165 A Case Of Rhabdomyolysis-Induced Hypocalcemia And Hypercalcemia | Case shows rhabdomyolysis causing simultaneous hypocalcemia and hypercalcemia; managed with calcium replacement in patient during recovery. | PMID:11454077 |

###### PMC – 10 Case Reports

**Table C.8:**
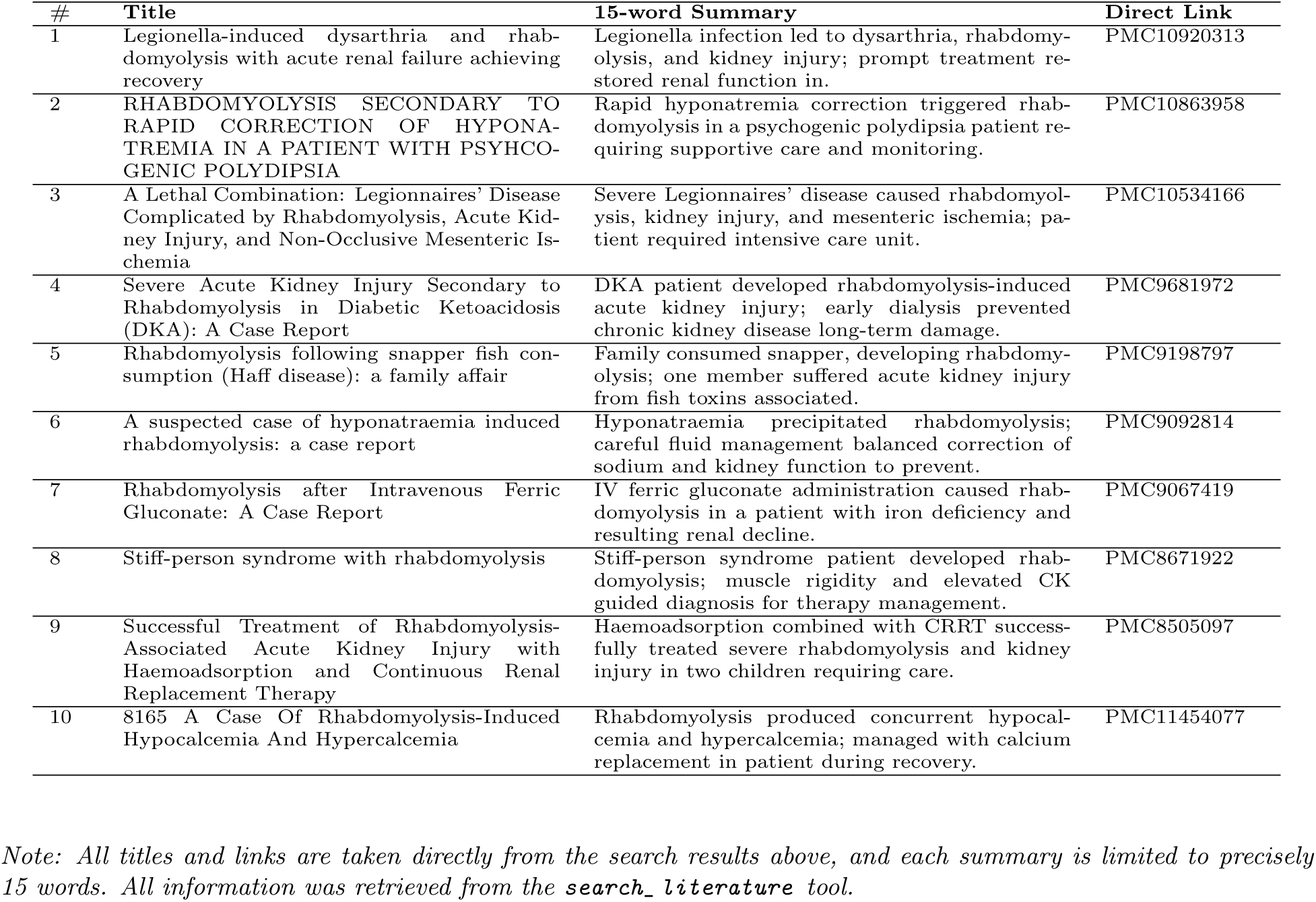
Case Reports Retrieved from PMC by gpt-oss-20b.

| # | Title | 15-word Summary | Direct Link |
| --- | --- | --- | --- |
| 1 | Legionella-induced dysarthria and rhabdomyolysis with acute renal failure achieving recovery | Legionella infection led to dysarthria, rhabdomyolysis, and kidney injury; prompt treatment restored renal function in. | PMC10920313 |
| 2 | RHABDOMYOLYSIS SECONDARY TO RAPID CORRECTION OF HYPONATREMIA IN A PATIENT WITH PSYCHOGENIC POLYDIPSIA | Rapid hyponatremia correction triggered rhabdomyolysis in a psychogenic polydipsia patient requiring supportive care and monitoring. | PMC10863958 |
| 3 | A Lethal Combination: Legionnaires' Disease Complicated by Rhabdomyolysis, Acute Kidney Injury, and Non-Occlusive Mesenteric Ischemia | Severe Legionnaires' disease caused rhabdomyolysis, kidney injury, and mesenteric ischemia; patient required intensive care unit. | PMC10534166 |
| 4 | Severe Acute Kidney Injury Secondary to Rhabdomyolysis in Diabetic Ketoacidosis (DKA): A Case Report | DKA patient developed rhabdomyolysis-induced acute kidney injury; early dialysis prevented chronic kidney disease long-term damage. | PMC9681972 |
| 5 | Rhabdomyolysis following snapper fish consumption (Haff disease): a family affair | Family consumed snapper, developing rhabdomyolysis; one member suffered acute kidney injury from fish toxins associated. | PMC9198797 |
| 6 | A suspected case of hyponatraemia induced rhabdomyolysis: a case report | Hyponatraemia precipitated rhabdomyolysis; careful fluid management balanced correction of sodium and kidney function to prevent. | PMC9092814 |
| 7 | Rhabdomyolysis after Intravenous Ferric Gluconate: A Case Report | IV ferric gluconate administration caused rhabdomyolysis in a patient with iron deficiency and resulting renal decline. | PMC9067419 |
| 8 | Stiff-person syndrome with rhabdomyolysis | Stiff-person syndrome patient developed rhabdomyolysis; muscle rigidity and elevated CK guided diagnosis for therapy management. | PMC8671922 |
| 9 | Successful Treatment of Rhabdomyolysis-Associated Acute Kidney Injury with Haemoadsorption and Continuous Renal Replacement Therapy | Haemoadsorption combined with CRRT successfully treated severe rhabdomyolysis and kidney injury in two children requiring care. | PMC8505097 |
| 10 | 8165 A Case Of Rhabdomyolysis-Induced Hypocalcemia And Hypercalcemia | Rhabdomyolysis produced concurrent hypocalcemia and hypercalcemia; managed with calcium replacement in patient during recovery. | PMC11454077 |

*Note: All titles and links are taken directly from the search results above, and each summary is limited to precisely 15 words. All information was retrieved from the `search_literature` tool.*

